# Common, low-frequency, rare, and ultra-rare coding variants contribute to COVID-19 severity

**DOI:** 10.1101/2021.09.03.21262611

**Authors:** Chiara Fallerini, Nicola Picchiotti, Margherita Baldassarri, Kristina Zguro, Sergio Daga, Francesca Fava, Elisa Benetti, Sara Amitrano, Mirella Bruttini, Maria Palmieri, Susanna Croci, Mirjam Lista, Giada Beligni, Floriana Valentino, Ilaria Meloni, Marco Tanfoni, Francesca Colombo, Enrico Cabri, Maddalena Fratelli, Chiara Gabbi, Stefania Mantovani, Elisa Frullanti, Marco Gori, Francis P. Crawley, Guillaume Butler-Laporte, Brent Richards, Hugo Zeberg, Miklos Lipcsey, Michael Hultstrom, Kerstin U. Ludwig, Eva C. Schulte, Erola Pairo-Castineira, John Kenneth Baillie, Axel Schmidt, Robert Frithiof, WES/WGS working group within the HGI, GenOMICC Consortium, GEN-COVID Multicenter Study, Francesca Mari, Alessandra Renieri, Simone Furini

**Affiliations:** Med Biotech Hub and Competence Center, Department of Medical Biotechnologies, University of Siena, Italy; Medical Genetics, University of Siena, Italy; University of Siena, DIISM-SAILAB, Siena, Italy; Department of Mathematics, University of Pavia, Pavia, Italy; Genetica Medica, Azienda Ospedaliero-Universitaria Senese, Italy; Istituto di Tecnologie Biomediche – Consiglio Nazionale delle Ricerche, Segrate (MI), Italy; Pharmacogenomics Unit, Istituto di Ricerche Farmacologiche Mario Negri IRCCS, Milan, Italy; Department of Biosciences and Nutrition, Karolinska Institutet, Stockholm, Sweden; Department of Medicine, Clinical Immunology and Infectious Diseases, Fondazione IRCCS Policlinico San Matteo, Pavia, Italy; Models and Algorithms for Artificial Intelligence (MAASAI) Research Group, Université Côte d’Azur, Inria, CNRS, I3S, France; Good Clinical Practice Alliance-Europe (GCPA) and Strategic Initiative for Developing Capacity in Ethical Review-Europe (SIDCER), Brussels, Belgium; Lady Davis Institute, Jewish General Hospital, McGill University, Montréal, Québec, Canada; Department of Epidemiology, Biostatistics and Occupational Health, McGill University, Montréal, Québec, Canada; Department of Human Genetics, McGill University, Montréal, Québec, Canada; Department of Twin Research, King’s College London, London, United Kingdom; Department of Neuroscience, Karolinska Institutet, Stockholm, Sweden; Anaesthesiology and Intensive Care Medicine, Department of Surgical Sciences, Uppsala University, Uppsala, Sweden; Hedenstierna Laboratory, CIRRUS, Anaesthesiology and Intensive Care Medicine, Department of Surgical Sciences, Uppsala University, Uppsala, Sweden; Integrative Physiology, Department of Medical Cell Biology, Uppsala University, Uppsala, Sweden; Institute of Human Genetics, University of Bonn, School of Medicine & University Hospital Bonn, Bonn, Germany; Institute of Psychiatric Phenomics and Genomics (IPPG), University Hospital, LMU Munich, Munich, 80336, Germany; Department of Psychiatry and Psychotherapy, University Hospital, LMU Munich, Munich, 80336, Germany; Institute of Virology, Technical University Munich/Helmholtz Zentrum München, Munich, Germany; MRC Human Genetics Unit, Institute of Genetics and Molecular Medicine, University of Edinburgh, Western General Hospital, Crewe Road, Edinburgh, EH4 2XU, UK; Roslin Institute, University of Edinburgh, Easter Bush, Edinburgh, EH25 9RG, UK; Intensive Care Unit, Royal Infirmary of Edinburgh, 54 Little France Drive, Edinburgh, EH16 5SA, UK

**Keywords:** COVID-19, Host genetics, Integrative polygenic score, Genetic science modelling, pathway enrichment analysis

## Abstract

The combined impact of common and rare exonic variants in COVID-19 host genetics is currently insufficiently understood. Here, common and rare variants from whole exome sequencing data of about 4,000 SARS-CoV-2-positive individuals were used to define an interpretable machine learning model for predicting COVID-19 severity. Firstly, variants were converted into separate sets of Boolean features, depending on the absence or the presence of variants in each gene. An ensemble of LASSO logistic regression models was used to identify the most informative Boolean features with respect to the genetic bases of severity. The Boolean features selected by these logistic models were combined into an Integrated PolyGenic Score that offers a synthetic and interpretable index for describing the contribution of host genetics in COVID-19 severity, as demonstrated through testing in several independent cohorts. Selected features belong to ultra-rare, rare, low-frequency, and common variants, including those in linkage disequilibrium with known GWAS loci. Noteworthly, around one quarter of the selected genes are sex-specific. Pathway analysis of the selected genes associated with COVID-19 severity reflected the multi-organ nature of the disease. The proposed model might provide useful information for developing diagnostics and therapeutics, while also being able to guide bedside disease management.

## Introduction

For almost two years COVID-19 has demonstrated itself to be a disease having a broad spectrum of clinical presentations: from asymptomatic patients to those with severe symptoms leading to death or persistent disease (“long COVID”) [1–3]. While developing vaccination programmes and other preventive measures to significantly dampen infection transmission and reduce disease expression, a much deeper and more precise understanding of the interplay between SARS-CoV-2 and host genetics is required to support the development of treatments for new virus variants as they arise. Furthermore, advances in modelling the interplay between SARS-CoV-2 and host genetics hold significant promise for addressing other complex diseases. In this study, we demonstrate the value of genetic modelling with its direct translatability into drug development and clinical care in the context of a severe public health crisis.

The identification of host genetic factors modifying disease susceptibility and/or disease severity has the potential to reveal the biological basis of disease susceptibility and outcome as well as to subsequently contribute to treatment amelioration [4]. From a scientific point of view, COVID-19 represents a particularly interesting and accessible complex disorder for modeling host genetic data because the environmental factor (SARS-CoV-2) can be readily identified by a PCR-based swab test. The still moderate viral genome variability has thus far been shown to have relatively low impact on disease severity [5] where currently age, sex, and comorbidities are the major factors predicting disease susceptibility and outcome [6]. While these factors certainly have significant value for prediction, they provide limited insights into disease pathophysiology and are of limited relevance for drug development.

Common variants in the human genome affecting the susceptibility to SARS-CoV-2 infections and COVID-19 severity have been successfully identified by Genome-Wide Association Studies (GWASs) [7–9]. However, these variants only explain a small fraction of trait variability and, as it is well documented, GWASs are difficult to interpret because they often associate non-coding variants with phenotype; therefore the relevant genes need to be pinpointed by deeper follow-up analyses. In contrast, next-generation sequencing based studies have identified variants in a few genes related to innate immunity which can solely underlie rare severe forms of COVID-19 [10–13]. In these rare affected families, the predictivity is high as the susceptibility for severe COVID-19 follows Mendelian inheritance patterns. However, these patients represent only a small proportion of those severely affected by COVID-19. Taken together, genetic findings can currently only explain a limited proportion of COVID-19 susceptibility and severity, in spite of the relatively high predicted heritability of COVID-19 and COVID-19 symptoms [14]. A better and more holistic understanding of host genetics could support the development of more specific, or even targeted drugs and treatment interventions leading to less morbidity and mortality.

The Italian GEN-COVID Multicenter Study collected more than two thousand biospecimens and clinical data from SARS-CoV-2-positive individuals [15], and whole exome sequencing (WES) analysis contributed to the identification of rare variants [7] and common polymorphisms [16–18] associated with COVID-19 severity. In 2020, we started to investigate how common variants may combine with rare variants to determine COVID-19 severity in WES data using a first small cohort of hospitalized patients. This pilot analysis revealed that the combination of rare and common variants could potentially impact clinical outcome [19]. We then proposed a new *post-Mendelian model* for a genetic characterization of the disorder [20] based on an adapted Polygenic Risk Score (PRS) [21], called *Integrated PolyGenic Score* (IPGS). This allowed us to reach a more precise disease severity prediction than that based on sex and age alone. In this article, we substantially improve this *post-Mendalian model* to include ultra-rare and low-frequency variants while also demonstrating that IPGS significantly contributes to predictivity in combination with - as well as alongside - age and sex, and is able to extract patient-specific genes. The IPGS predictivity was also sustained in three independent European cohorts of the WES/Whole-Genome Sequencing study working group within the COVID-19 Host Genetics Initiative [22].

## Material and Methods

### Contributing cohorts

Five different cohorts (from Germany, Italy, Quebec, Sweden, UK) contributed to this study as described in **Supplementary Table 1**. For multi ancestry cohorts (Quebec and UK) the subpopulation of European Ancestry was included in the study. Institutional Review Board approval was obtained for each study (**Supplementary Table 1**).

### Phenotype definitions

The training of the model proposed for predicting the severity of COVID-19 requires as inputs the exome variants, age, sex, and COVID-19 severity assessed using a modified version of the WHO COVID-19 Outcome Scale [23] as coded into the following six classifications: 1. death; 2. hospitalized receiving invasive mechanical ventilation; 3. hospitalized, receiving continuous positive airway pressure (CPAP) or bilevel positive airway pressure (BiPAP) ventilation; 4. hospitalized, receiving low-flow supplemental oxygen; 5. hospitalized, not receiving supplemental oxygen; and 6. not hospitalized. The aim of the model is to predict a binary classification of patients into mild and severe cases, where a patient is considered severe if hospitalized and receiving any form of respiratory support (WHO severity-grading equal to 4 or higher in 8 points classification). The next section describes how the annotation of exome variants and the selection of patients were performed in the GEN-COVID cohort. Following this the training and testing of the model are described.

### Massive parallel sequencing

#### GEN-COVID cohort

Whole Exome Sequencing with at least 97% coverage at 20x was performed using the Illumina NovaSeq6000 System (Illumina, San Diego, CA, USA). Library preparation was performed using the Illumina Exome Panel (Illumina) according to the manufacturer’s protocol. Library enrichment was tested by qPCR, and the size distribution and concentration were determined using Agilent Bioanalyzer 2100 (Agilent Technologies, Santa Clara, CA, USA). The Novaseq6000 System (Illumina) was used for DNA sequencing through 150 bp paired-end reads. Variant calling was performed according to the GATK4[24] best practice guidelines, using BWA [25] for mapping and ANNOVAR [26] for annotating.

#### Swedish cohort

Whole Exome Sequencing was performed using the Twist Bioscience exome capture probe and was sequenced on the Illumina NovaSeq6000 platform. Data were then analyzed using the McGill Genome Center bioinformatics pipeline (https://doi.org/10.1093/gigascience/giz037) in accordance with GATK best practices.

#### DeCOI Germany

800-1000 ng of genomic DNA of each individual was fragmented to an average length of 350 bp. Library preparation was performed using the TruSeq DNA PCR-free kit (Illumina, San Diego, CA, USA) according to the manufacturer’s protocol. Whole genome sequences were obtained as 150 bp paired-end reads on S4 flow cells using the NovaSeq6000 system (Illumina). The intended average sequencing depth was 30X. The DRAGEN pipeline (Illumina, version 3.6.3 or 3.5.7) was used for alignment and joint variant calling was performed with the Glnexus software (version 1.3.2). Individuals with a 20-fold coverage in less than 96% of the protein coding sequence were removed as well as related individuals to retain only from related pairs. Variant QC was performed using hail (version 0.2.58). European individuals were selected by performing PCA analysis along with the 1000 genomes data. Finally, annotation was performed using Variant Effect Predictor (VEP, version 101).

#### BQC-19

Whole genome sequencing at mean coverage of 30x was performed on the Illumina NovaSeq6000 platform, then analyzed using the McGill Genome Center bioinformatics pipeline (https://doi.org/10.1093/gigascience/giz037), in accordance with GATK best practice guidelines.

#### GenOMICC/ISARIC4C

Whole genome sequencing at mean coverage of 20x was performed on the Illumina NovaSeq6000 platform and then analysed using the Dragen pipeline (software v01.011.269.3.2.22, hardware v01.011.269) . Variants were genotyped with the GATK GenotypeGVCFs tool v4.1.8.1.

### PC analysis

The standard analysis of Principal Components was performed and the first principal components turned out to be connected with the patient’s ethnicity collected in the medical records. Therefore, the genetic ancestry of the patients was estimated using a random forest classifier trained on samples from the 1000 genomes project and using as input features the first 20 principal components computed from the common variants by PLINK [27]. In order to avoid bias in the analysis due to the different ethnicity, only patients of genetic European ancestry were retained for further analyses.

### Definition of the Boolean features

Variants were converted into 12 sets of Boolean features to better represent the variability at the gene-level. Firstly, any variant not impacting on the protein sequence was discarded. Then the remaining variants were classified according to their minor allele frequency (MAF) as reported in gnomAD for the reference population as: ultra-rare, MAF<0.1%; rare, 0.1%<=MAF<1%; low-frequency, 1%<=MAF<5%; and common, MAF>=5%. Non-Finnish European (NFE) was used as a reference population. SNPs with MAF not available in gnomAD were treated as ultra-rare. INDELs with frequency not available in gnomAD were treated as ultra-rare when present only once in the cohort and otherwise discarded as possible artefacts of sequencing. For the ultra-rare variants, 3 alternative Boolean representations were defined, which are designed to capture the autosomal dominant (AD), autosomal recessive (AR), and X-linked (XL) model of inheritance, respectively. The AD and AR representations included a feature for all the genes on autosomes. These features were equal to 1 when the corresponding gene presented at least 1 for the AD model, or 2 for the AR model, and variants in the ultra-rare frequency range and 0 otherwise. The XL representation included only genes belonging to the X chromosome. These features were equal to 1 when the corresponding gene presented at least 1 variant in the ultra-rare frequency range and 0 otherwise. The same approach was used to define AD, AR, and XL Boolean features for the rare and low-frequency variants. Common variants were represented using a different approach that is designed to better capture the presence of alternative haplotypes. For each gene, all the possible combinations of common variants were computed. For instance, in the case of a gene belonging to an autosome with 2 common variants (named A and B), 3 combinations are possible (A, B, and AB), and (consequently) 3 Boolean features were defined both for the AD and AR model. In the AR model each of these 3 features was equal to 1 if all the variants in that particular combination were present in the homozygous state and 0 otherwise. The same rule was used for the AD model, but setting the feature to 1 even if the variants in that particular combination are in the heterozygous state. In both models, AD and AR, a further feature was defined for each gene to represent the absence of any of the previously defined combinations. In the AD model this feature was equal to 1 if no common variant is present and 0 otherwise; in the AR model, it is equal to 1 if no common variant is present in the homozygous state and 0 otherwise. The same approach was used to define the set of Boolean features for common variants in genes belonging to the X chromosome. The full list of Boolean representations is reported in **Supplementary Table 2**.

### Model Training

The dataset was divided into a training set and a testing set (90/10), and the entire procedure described in this section was performed using only samples in the training set. A bootstrap approach with 100 iterations was adopted to train the model. At each bootstrap iteration, 90% of the samples were selected (without replication), and the following two steps were performed: (step 1) selection of the most relevant features for each Boolean representations; and (step 2) definition of the weights of the *Integrated Polygenic Score* (*IPGS*). After the 100 bootstrap iterations, the information extracted on relevant features and weighting factors are merged to define the final *IPGS* (step 3). The *IPGS* is then used, together with age and sex, for training a model that predicts the COVID-19 severity (step 4). These 4 steps of the training procedure are described in detail in the next subsections. Because the model is based on a combination of rare and common variants, the training procedure should be performed using a dataset with homogeneous ancestry.

#### Step 1: Features’ Selection

The subsets of the most relevant features were identified using logistic regression models with Least Absolute Shrinkage and Selection Operator (LASSO) regularization. Separate logistic models were trained for each of the 12 sets of Boolean features. The predicted outcome variable for each of these models was a re-classified phenotype adjusted by age and sex. In order to compute these re-classified phenotypes as adjusted by age and sex, the patients were first divided into males and females. Then for each sex an ordinal logistic regression model was fitted by using the age to predict the WHO phenotype classification into 6 grades. The ordinal logistic regression model was chosen as: it imposes a simple monotone relation between input feature and target variable; and it provides easily interpretable thresholds between the predicted classes. The patients with a predicted grading equal to the actual grading were excluded. The remaining patients were divided into two classes depending on whether their actual phenotype was milder or more severe than the one expected for a patient of that age and sex. This procedure has the benefit of isolating patients whose genetic factors are most important for predicting COVID-19 severity. This binary trait, i.e. phenotype more/less severe than expected, was used as the outcome variable for the 12 LASSO logistic models based on the 12 separate Boolean representations. For each LASSO model, the regularization strength was optimized by 10-folds cross-validation with 50 equally spaced values in the logarithmic scale in the range [10^−2^, 10^1^]. The optimal regularization strength was selected as the one with the best trade-off between the simplicity of the model and the cross-validation score, i.e. as the highest regularization strength providing an average score closer to the highest average score than 0.5 standard deviations. Once the regularization strength was defined, the LASSO model was re-trained using all the samples in that particular bootstrap iteration. The features with non-null coefficients are the ones selected for the next step. In summary, for each bootstrap iteration, this procedure returns 12 lists of features (one for each Boolean representation) that are expected to be the most important features for predicting the phenotype adjusted by age and sex (in that particular bootstrap iteration).

#### Step 2: Weights of the Integrated Polygenic Score (IPGS)

In the previous step, the Boolean representations are considered isolated from each other. The aim of the *IPGS* is to combine information from different representations (**Equation 1**). In order to reach this goal, it is necessary to compute the relative weights of the different contributions. For each bootstrap iteration, the list of relevant features extracted as described in the previous section are used to compute the number of features that are associated with mildness or severity for the different frequency ranges. For instance, 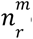 corresponds to the number of features associated with the mild phenotype coming from Boolean features computed for variants in the frequency range [0.1%, 1%]. A feature is considered associated with the mild phenotype when its coefficient in the LASSO model estimated in step 1 is negative, i.e. it contributes to the prediction of the phenotype adjusted by age and sex in the direction of a phenotype less severe than what expected at that particular age and sex. The same rule, applied to the corresponding Boolean representation, is used to define the other feature-counts appearing in **Equation 1**. The weighting factors in Equation 1 were estimated as the ones that maximize the Silhouette coefficient of the separation between the clusters of patients more/less severe than expected. The minimization was performed with weighting factors restricted to the following ranges: *F_LF_* ∈ [1, 4], *F_R_* ∈ [2, 8] and *F_UR_* ∈ [5, 100]. This procedure returns 3 optimal values for the weighing factors associated with each bootstrap iteration.

#### Step 3: *IPGS* definition

In this step, the data extracted at each bootstrap iteration in steps 1 and 2 are combined to define the *IPGS*. Firstly, for each of the Boolean features, of all the 12 representations, the number of times this feature was selected in the 100 bootstrap iterations is computed. Then, the entire bootstrap procedure is repeated using random input phenotypes, and the 5^th^ percentile of the number of times that a feature is associated with a random phenotype is estimated. This threshold, computed separately for each Boolean representation, was used to select which Boolean features are included in the final model. As no significant association is expected among the Boolean features and the random phenotype, the threshold of the 5^th^ percentile is expected to exclude with a 95% level of confidence the possible false positive associations. For the GEN-COVID cohort, 7249 features were selected by this procedure, which correspond to ∼4.4% of the initial number of Boolean features. The weighing factors in Equation 1 were computed as the median values of the estimates obtained in the 100 bootstrap iterations.

#### Step 4: Training of the predictive model based on age, sex, and *IPGS*

The procedure described in the previous sections completely defines how to calculate the *IPGS*. The predictive model of the binary COVID-19 severity (hospitalized patients with any form of respiratory support versus all other patients) was defined as a logistic model that uses as input features IPGS, age, and sex. It should be noted that in steps 1-3, only patients that deviates from their expected severity based on age and sex were used. The procedure was designed to isolate the genetic basis of COVID-19 severity. Instead, in this final step, *IPGS*, age and sex are combined to predict the actual COVID-19 severity, i.e. hospitalized patients with any form of respiratory support. In order to prevent overfitting, the model was fitted using 466 samples different from the training set adopted in steps 1-3. During the fitting procedure, the class unbalancing is tackled by penalizing the misclassification of the minority class with a multiplicative factor inversely proportional to the class frequencies. The percentile normalization of the *IPGS* scores is performed within each cohort. An alternative logistic model that used as input features only age and sex was also fitted on the same training set. The comparison between the two models is intended to evaluate if the genetic information summarized in the *IPGS* improves the prediction of severity compared to a model based on age and sex alone. A further logistic regression model is fitted by only considering the *IPGS* variable.

### Model Testing

The training procedure returned 2 logistic models to be compared: one using as input features only age and sex, and the other one using as input features age, sex, and the *IPGS*. These models were tested, without any further adjustment, using other cohorts of European ancestry. The performances of the two models, with and without *IPGS*, were evaluated and compared in terms of accuracy, precision, sensitivity, and specificity. The increases of the performances are evaluated with respect to the performances of a model where the values of the *IPGS* feature have been shuffled, by computing the p-value on the empirical null distribution. In addition, the empirical probability density function of *IPGS* has been estimated for the severe and non-severe patients of the cohort including both train and test sets and a t-test is carried out to evaluate whether the means of the two distributions were significantly different. As a further evaluation of the importance of the *IPGS* score on the severity prediction, univariate logistic regression models using as independent variables age (continuous represented in decades), sex, and *IPGS* were fitted to the dataset that combines both the training set and the testing sets for a total of 2,240 patients. These models were used to estimate the odds ratios and the p-values of the association with the severe phenotype. Furthermore, a multivariable logistic regression was fitted using IPGS, age, and sex together. Finally, a multivariable logistic regression was performed using as predictor variables: IPGS, age, sex and comoribidities (congestive/ischemic heart failure; asthma/COPD/OSAS; diabetes; hypertension; cancer). This latter model has been fitted in the training set, where the information on comorbidities was available.

### Pathway analysis

Pathway analysis was made using a ranked GSEA approach [28–29], modified according to the specificity of our data. The metrics for gene ranking was calculated on the basis of the results of feature selection models, weighting in each Boolean feature both beta values and the number of bootstrap iterations where it was found significantly associated with severity/mildness (**Supplementary Tables 3-6**). All the Boolean features that were found significant in at least one of the models were included in the list. As the sign of beta depends on which allele is taken as reference (which is relative for common variants), we decided to use absolute beta values for all the features. To also weight the importance according to variant frequency, we used the F values from the *IPGS* score for the four categories (Ultra-rare 5, Rare 4, Low Frequency 2, Common 1). Finally, we summed all the weights of each Boolean feature by gene. Briefly:

W_geneA_ = ∑_FeaturesGeneA_ ABS(meanβ)∗count∗F

Pathway enrichment analysis was made using the GSEA-preranked module (v. 7.2.4) of the Genepattern platform [26], on several pathway categories (BIOCARTA, KEGG, REACTOME, GOBP, HALLMARKS, C7 and C8), limiting the size of genesets to the 10-300 range and performing 10,000 permutations. The networks showing similarity of significant pathways were built using the EnrichmentMap algorithm [30] in the Cytoscape suite (v. 3.8.2) [31–32]. Parameters used for network creations are: Jaccard Overlap Combined Index (k constant = 0.5), edge cutoff 0.05.

### Website and data distribution

The coordination of international partners has been possible through the Host Genetics Initiative (HGI) (https://www.covid19hg.org/projects/).

Results can be shared through the Gen-Covid website (https://sites.google.com/dbm.unisi.it/gen-covid).

## Code availability

Data analyses were performed using Python with the Scipy ecosystem [33], and the scikit-learn library [34]. Statistical association was done with the statsmodel Python library. The code is freely available at the github repository: https://github.com/gen-covid/pmm.

## Results

### The post-Mendelian paradigm for COVID-19 modelization for combining interpretability with predictivity based on ultra-rare, rare, low-frequency, and common variants

The aim of the present study was to develop an easily interpretable model that could be used to predict the severity of COVID-19 from host genetic data. Patients were considered severe when hospitalized and receiving any form of respiratory support. The focus on this target variable is motivated by the practical importance of rapidly identifying which patients are more likely to require oxygen support, in an effort to prevent further complications. Interpretability has been a guiding principle in the definition of the machine learning model, as only a readily interpretable model can provide useful and reliable information for clinical practice while also contributing significantly to diagnostic, and therapeutic targeting. The high dimensionality of host genetic data poses a serious challenge to evident and reliable interpretability. So far, the development of a robust predictive model able to make a direct association between single variants and disease severity grading based on an accurate analysis of the vast number of host genetic variants compared to a much smaller number of individual patients has proven to be too complex and ultimately unreliable. In order to address the complexity with predictive reliability, an enriched gene-level representation of host genetic data was modeled in a machine learning framework. The complexity of COVID-19 immediately suggests that both common and rare variants are expected to contribute to the likelihood of developing a severe form of the disease. However, the contribution of common and rare variants to the severe phenotype is not expected to be the same. A single rare variant that impairs the protein function might cause a severe phenotype by itself after viral infection, while this is not so probable for a common polymorphism, which is likely to have a less marked effect on protein functionality. These observations led to the definition of a score, named *IPGS*, that includes data regarding the variants at different frequencies:

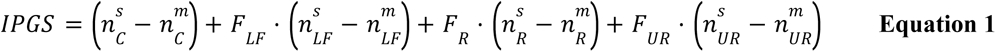

In **Equation 1**, *n* variables are used to indicate the number of input features of the predictive model that promote the severe outcome (superscript *s*) or that protect from a severe outcome (superscript *m*) and with genetic variants having Minor Allele Frequency (MAF)>=5% (common, subscript *C*), 1%<MAF<= 5% (low-frequency, subscript *LF*), 0.1%<MAF<=1% (rare, subscript *R*), and MAF<0.1% (ultra-rare, substript *UR*). The features promoting or preventing severity were identified by an ensemble of logistic models, as described in the next section. The weighting factors *F_LF_*, *F_R_*, and *F_UR_* were included to model the different penetrant effects of low-frequency, rare, and ultra-rare variants, compared to common variants. Thus, the 4 terms of **Equation 1** can be interpreted as the contributions of common, low-frequency, rare, and ultra-rare variants to a score that represents the genetic propensity of a patient to develop a severe form of COVID-19.

### Feature selection and gene discovery

The definition of the single terms of the *IPGS* formula requires 4 separate steps (**Fig. 1**): (1) the definition of a severity phenotype adjusted by age and sex; (2) the conversion of genetic variants into Boolean features that represent the presence of variants in different frequency ranges in each gene; (3) the selection of those features that are associated with disease severity; and (4) the optimization of the weighting factors appearing in Equation 1. These 4 steps were executed using data from a training set extracted from the GEN-COVID dataset (90% of the patients, n=1780, see Methods). The phenotype adjusted by age and sex was computed using an ordered logistic regression model, with the pourpose of facilitating the extraction of features associated with the genetic basis of COVID-19 severity (**Fig. 1B**). The conversion of genetic variants into Boolean features led to the definition of 12 separate sets of input features (**Supplementary Table 2**). The set of input features “ultra-rare_autosomal dominant” (UR_AD) is designed to represent in a binary way an autosomal dominant hereditary model associated with variants with MAF lower than 0.1%; i.e. these Boolean features are equal to 1 for genes presenting at least one variant in this frequency range. Similarly, the set of input features “ultra-rare_autosomal recessive” (UR_AR) and “ultra-rare_X-linked” (UR_X) were designed to describe the autosomal recessive and X-linked models of inheritance of ultra-rare variants. Analogous principles were used for rare and low-frequency variants. In the case of common variants, the same 3 sets of Boolean features representing the autosomal dominant, autosomal recessive, and X-linked models of inheritance were used. However, instead of simply defining the binary variables as “absence/presence of variants”, the absence/presence of variant combinations was tested (**Fig. 1C**).

**Figure 1.**
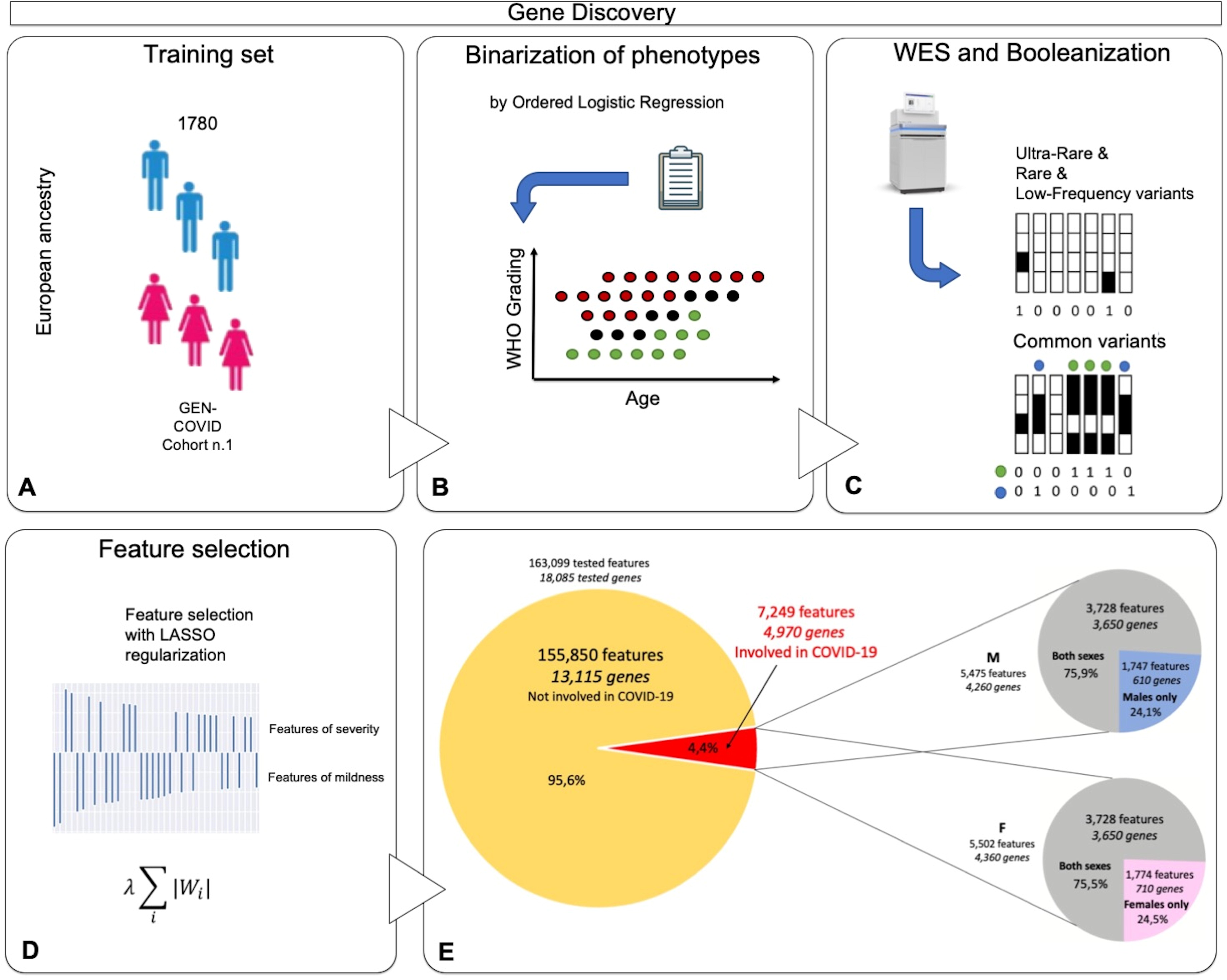
Feature selection and gene discovery. **A)** Whole Exome Sequencing (WES) data stored in the Genetic Data Repository of the GEN-COVID Multicenter Study (GCGDR) and coming from biospecimens of 1,780 SARS-CoV-2 PCR positive subjects of European ancestry of different severity were used as the training set. **B)** Clinical severity classification into severe and mild cases was performed by Ordered Logistic Regression (OLR) starting from the WHO grading and patient age classifications. **C)** WES data were binarized into 0 or 1 depending on the absence (0) or the presence (1) of variants (or the combination of two or more variants only for common polymorphisms) in each gene. **D)** LASSO logistic regression feature selection methodology on multiple train-test splits of the cohort leads to the identification of the final set of features contributing to the clinical variability of COVID-19 **(E)**. From the initial 163,099 cumulative features (divided into 36,540 ultra rare, 23,470 rare, 13,056 low frequency and 90,033 common features) in 12 Boolean representations, the selected features contributing to COVID-19 clinical variability are 7,249 and they are reported in the **Supplementary Tables 3-6**. The total number of genes contributing to COVID-19 clinical variability was 4,260 in males and 4,360 in females, 75% of which were in common.

The Boolean representation of the genetic variability described in the previous paragraph significantly reduces the dimensionality of the problem. However, the number of input features is still orders of magnitude higher than the number of patients that can be reasonably collected for training the model. In order to reduce the number of input features, a feature selection strategy based on logistic models with the validated **L**east **A**bsolute **S**hrinkage and **S**election **O**perator (LASSO) regularization was employed (**Fig. 1D**). The aim of LASSO regularization is to minimize (shrink) the number of coefficients of the model, consequently minimizing the number of input features used for predicting outcomes. Separate logistic models with LASSO regularization were trained for the 12 sets of Boolean features for predicting COVID-19 severity, allowing us to identify the relevant features for each set. About 4% of the cumulative tested features were found to contribute to COVID-19 variability in severity (**Fig. 1D**).

### Biological interpretability of extracted genetic features

Selected genes contributed by ultra-rare, rare, low-frequency variants, or/and common variants (**Fig. 2A-D** and **Supplementary Tables 3a-g**). Specifically, 54% contributed by only one, 29% by two, 11% by three, and 6% by four types of variants. Around 25% of the genes were sex-specific. The latter group includes either genes located on the X chromosome, such as *TLR7* and *TLR8* in males, or genes regulated in opposite directions by androgens and estrogens when contributing with less penetrant common variants, such as p.L412F in *TLR3* and p.D603N in *SELP* gene (**Fig. 2A-D**).

**Figure 2.**
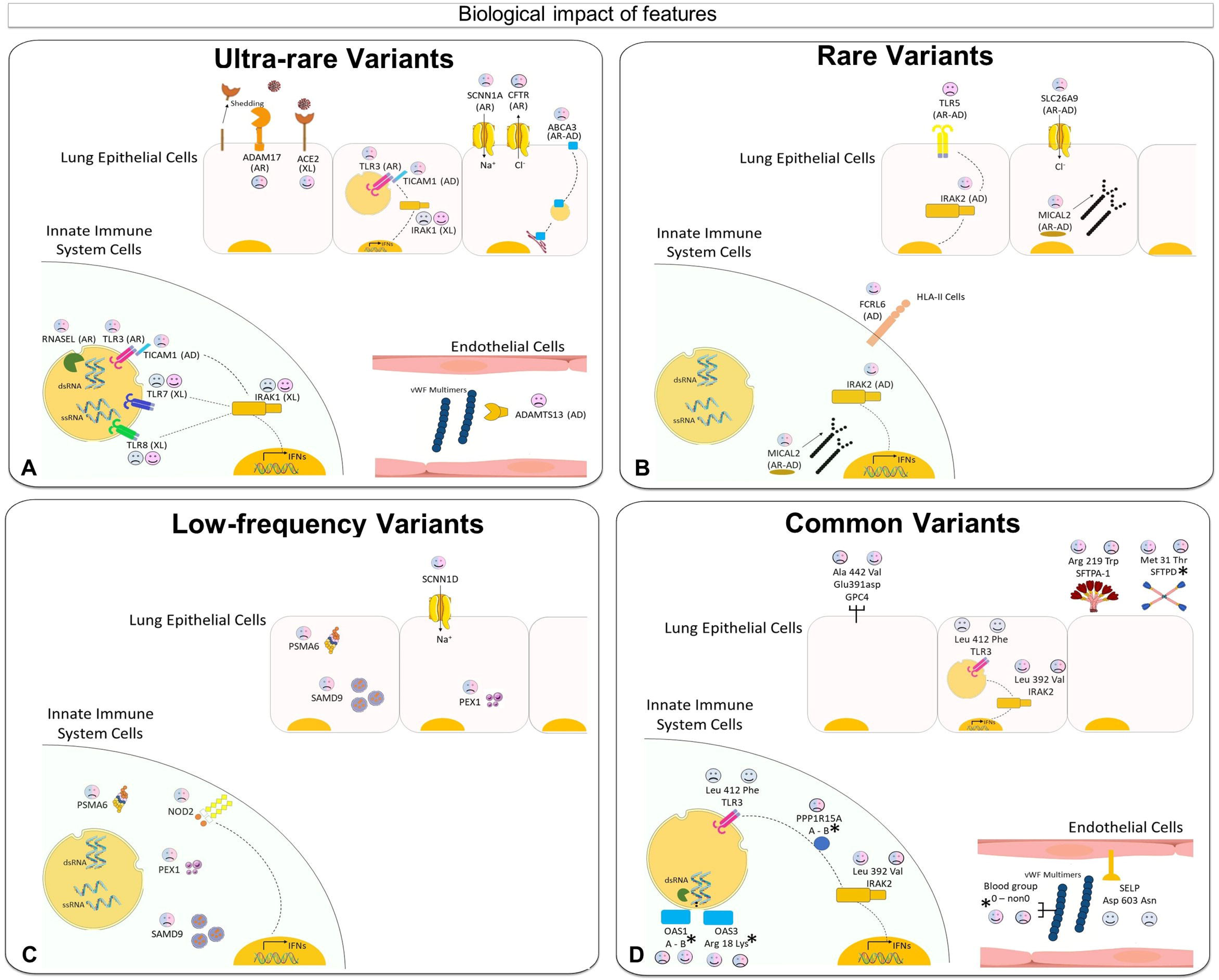
Biological impact of ultra-rare, rare, low-frequency, and common features. Examples of ultra-rare **(A)**, rare **(B)**, low-frequency **(C)**, and common **(D)** features are illustrated in panel A-D. The complete list of features is presented in **Supplementary table 3-6**. 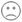 =contributing to COVID-19 severity; 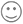 = contributing to COVID-19 mildness. Pink faces= contributing to females only; Blue faces = contributing to males only; Pink/Blue faces = contribution in both sexes. In parentheses: AD=autosomal dominant inheritance; AR=autosomal recessive inheritance; XL=X-linked recessive inheritance. **A)** ultra-rare mutations in the RNA sensor *TLR7*, *TLR3*, and *TICAM1* (encoding TRIF protein), already reported associated with XL, AR and AD inheritance [10–13] impair interferon (IFNs) production in innate immune system cells. Mutations in *TLR8*, as well as of the signal transducer *IRAK1* also impair interferon production. The specific location of *TLR7/8* and *IRAK1* (on the X chromosome) as well as X-inactivation escaping are responsible for opposite effects in males and females. Mutation in *RNASEL* impair the antiviral effect of the gene. In lung epithelial cells, *ACE2* ultra-rare variants (on the X chromosome) exert protective effects (probably) due to lowering virus entrance, while ultra-rare variants in *ADAM17* (might) reduce the shedding of *ACE2* and induce a severe outcome. The same is true for *CFTR* and *SCNN1A* (encoding ENaCA protein and involved in a CFTR-related physiological pathway), and the lipid transporter *ABCA3*[44]. Mutations of *ADAMTS13* in vessels reduce the cleavage of the multimeric von Willebrand Factor (VWF), leading to thrombosis; **B)** Rare variants of the estrogen regulated *TLR5* are associated with severity in females. Rare variants of the CFTR-related *SLC26A9* are associated with severity in both sexes. This ion transporter has three discrete physiological modes: nCl(-)-HCO(3)(-) exchanger, Cl(-) channel, and Na(+)-anion cotransporter. Other examples of rare mutations associated with severity are the NK and T cell receptor *FCRL6*, IFN signal transducer *IRAK2*, and the actin depolymerization *MICAL2*; **C)** low-frequency variants in another CFTR-related gene, *SCNN1D* (encoding for ENaCD protein) are associated with mildness, while rare variants in the following genes are associated with severity: cargo protein SPMA6, vesicle formation PEX1, inflammatory protein *NOD2* (*CARD15*); **D)** A number of coding polymorphisms, indicated with an asterisk, are in LD with genomic SNPs already associated with COVID-19 (The complete list is presented in **Supplementary Table 11**) [8, 37]. In some cases, such as the case of *SFTDP*, the genomic SNP is the coding polymorphism itself. Of note are the genes of surfactant proteins associated with severe disease: *SFTDP* gene encoding for SP-D protein and *SFTPA1* gene encoding for SP-A protein; the signal transducer, *PPP1R15A* gene encoding for GADD34 protein. OAS1 and OAS3 related to RNA clearance of *RNASEL* (reported in panel A as having ultra-rare mutations; included here should also be the already reported *TLR3412* [17]; the already reported SELP603 related to thrombosis [18]. Note: *OAS1* haplotype A= c.1039-1G>A, (p.(Gly162Ser)), (p.(Ala352Thr)), (p.(Arg361Thr)), (p.(Gly397Arg)), (p.(Thr358Profs*26)). *OAS1* haplotype B = haplotype without the variant combination in haplotype A.

Among the extracted ultra-rare variants there was a group of genes, such as *TLR3*, *TLR7* and *TICAM1*, already shown to be directly involved in the Mendelian-like forms of COVID-19 (**Fig. 2A** and **Supplementary Table 3a-b**). Furthermore, another group of genes are natural candidates because of their function: these include the *ACE2* shedding protein *ADAM17*, *CFTR*-related genes, genes involved in glycolipid metabolism, genes expressed by cells of the innate immune system, and genes involved in the coagulation pathway. Finally, a group of genes led by *ACE2* (if affected by ultra-rare variants) confers protection from the severe disease. This group includes several genes whose mutations are responsible for auto-inflammatory disorders.

Among the rare variants extracted, we identified some genes as candidates for COVID-19 severity, including *TLR5* and *SLC26A9* as well as other genes involved in the inflammatory response **Fig. 2B** and **Supplemnetary Table 4a-b**).

Among the low-frequency variants extracted, we identified some genes associated with either severity or protection from severe COVID-19 that are linked to the *CFTR* pathway (e.g., *PSMA6*) as well as specific genes involved in the immune response (e.g., *NOD2*) (**Fig. 2C** and **Supplementary Table 5a-b**).

The model was also able to identify a group of extracted common variants already shown to be linked to either severe or mild COVID-19 (**Fig. 2D** and **Supplementary Table 6a-b**). Among them are the L412F *TLR3* and D603N *SELP* polymorphisms, already reported to be associated with the severe disease [17–18] and several coding polymorphisms in Linkage Disequilibrium (LD) with already reported genomic SNP, such as the ABO blood group, *OAS1*-*3* genes, *PPP1R15A* gene and others [4]. In conclusion, considering their functions, genes involved in the immune and inflammatory responses, or those involved in the coagulation pathway and NK and T cell receptor, are to be considered natural candidates for severe or mild COVID-19.

### Integrated PolyGenic Score definition

The Boolean features selected by the LASSO logistic models were used to calculate the *n* variables in **Equation 1** (**Fig. 3A-B**). The corresponding weights (*F* variables) were defined by optimizing the separation between severe and mild cases as offered by the *IPGS* formula. The optimization was measured using the Silhouette coefficient, and the optimal values were computed using a grid-search approach over a predefined grid (*F_LF_* ∈ [1, 4], *F_R_* ∈ [2, 8], and *F_UR_* ∈ [5, 100]). This optimization returned values of **2, 4,** and **5** for the low-frequency, rare, and ultra-rare variants, respectively (**Fig. 3C-D**).

**Figure 3.**
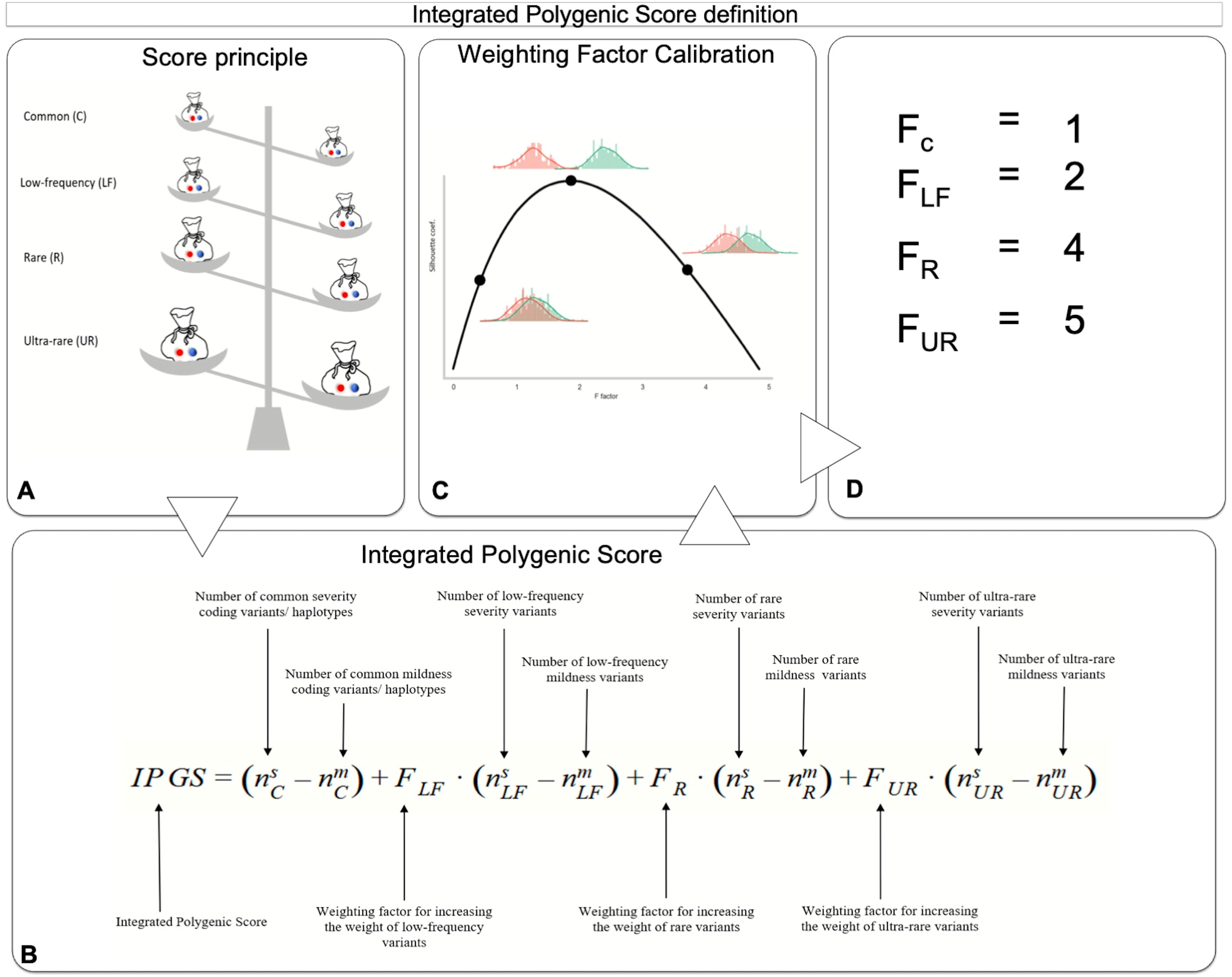
Integrated PolyGenic Score Definition. **A)** The model is based on the comparison of Boolean features of severity versus Boolean features of mildness. **B)** Graphic representation of the *IPGS* formula used for this model. **C)** Principle for the calibration of different weighting factors based on the separation of severe and mild cases. **D)** The obtained value for low-frequency, rare, and ultra-rare, being F=1 for common variants. Common variants are indicated as common haplotypes since they are intended as combinations of coding variants within a single gene (see Fig. 1C and the **Material and Methods** section).

### Pathway analysis

In order to understand the biological mechanisms underlying the variability of disease, we performed a pathways analysis of the genes carrying variants discovered in the feature selection described above. The features obtained with this approach do not have the same predicted impact and are not discovered with the same confidence. Therefore, we decided to perform a rank-based pathway analysis, with genes with the highest impact and confidence ranking highest in the list, rather than a simple over-representation approach. We ranked all the genes that were found to be significantly associated with severity/mildness in at least one bootstrap repetition, based on a score that takes into account three parameters: average coefficient in the LASSO models selecting the feature, number of significant bootstrap results, and the *F* correction factor for the frequency category used in the *IPGS* (detailed in the Methods section below). For genes with more than one significant Boolean feature, we summed up the scores of each feature. Gene Set Enrichment Analysis (GSEA) was then performed using two separate ranked gene lists (**Supplementary Table 7)** for females and males, followed by the generation of similarity networks using EnrichmentMap (Fig. 4A). The usage of rank-based search method allows to identify statistically significant pathways starting from extensive list genes, as each gene is associated with its specific importance. Although no pathways satisfied the 0.25% FDR threshold normally required for standard GSEA analyses, the set of pathways considered significant using more relaxed thresholds on p values were shown to group in meaningful modules, providing useful information on pathogenetic mechanisms and on the genes that could explain how they can be affected. The network of all the pathways significantly enriched in both females and males ranked gene lists (p<0.01, n=25) is depicted in **Fig. 4B**, while the network of all the pathways enriched in either females or males with a more stringent p-value (p<0.005, n=100) is shown in **Fig. 4C**. Detailed information on the names of the pathways and p-values of enrichment is reported in **Supplementary Figures 1 and 3**. For the most representative pathways of each network, the heatmaps of the genes with their weights of association to disease variability are shown in **Fig. 4D** and **Supplementary Figures 2 and 4**, while gene lists and gene weights for all the significant pathways are reported in **Supplementary Table 8.**

**Figure 4.**
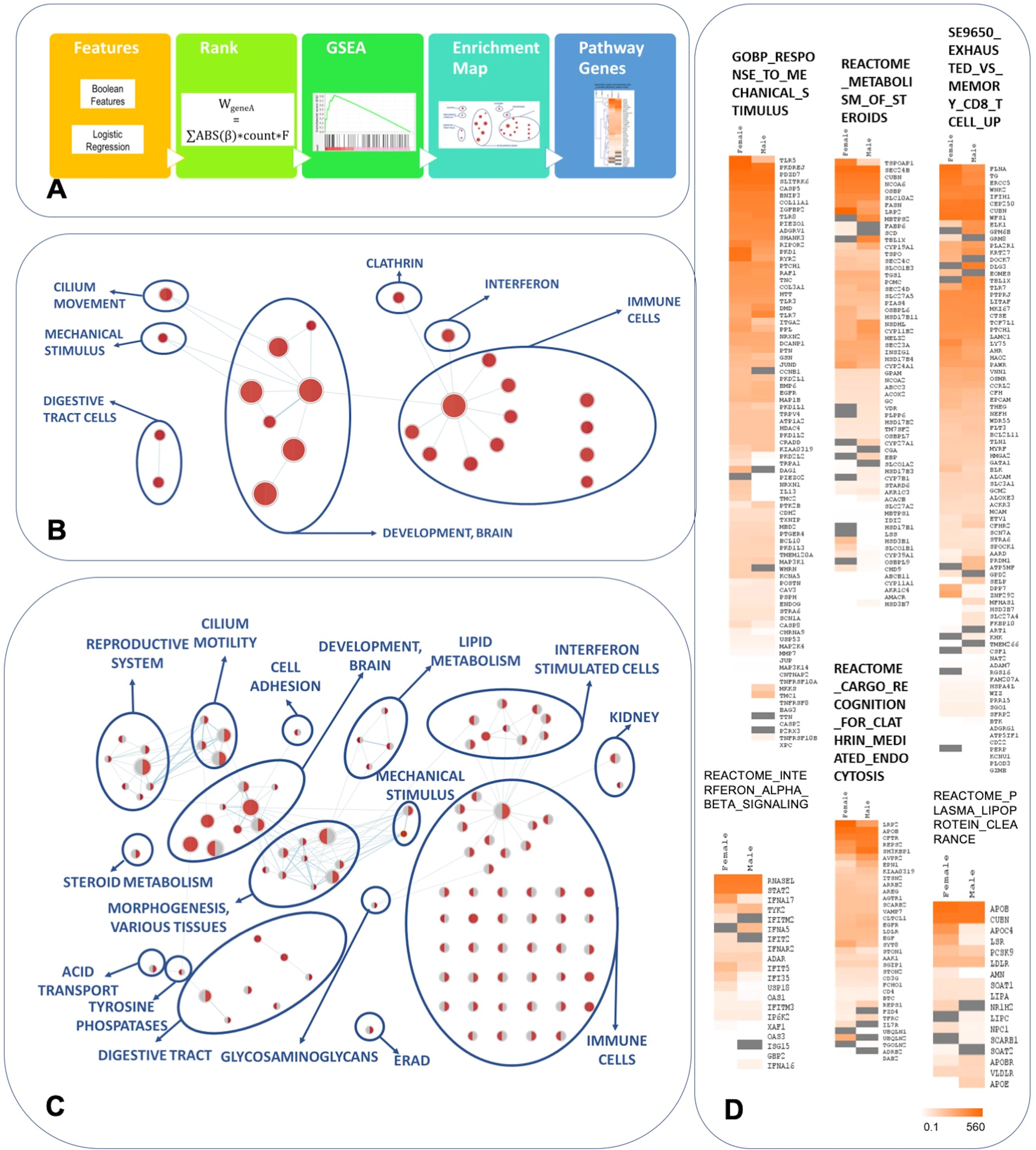
Pathway enrichment analysis of the genes associated with disease severity/mildness. **A)** Workflow of the analysis. Genes corresponding to Boolean features found to be associated at least once were ranked based on a composite score and subjected to Gene Set Enrichment Analysis. Two separate ranked gene lists for females (7,317 genes, weight range 3×10-5-561) and males (7,325 genes, weight range 7×10-5-452) were used. The list of significant pathways was analysed and presented as a similarity network: **B)** Similarity network of the pathways with a significant enrichment both in females and males (p<0.01). The size of the circles is proportional to the pahway size. Significance above threshold is indicated by the red color. **C)** Similarity network of the pathways with a significant enrichment either in females (red left half of the circles) or males (red right half of the circles) (p<0.005). **D)** Heatmaps of the genes belonging to a selection of pathways of interest. The color gradient represents the weight of each gene, calculated and described in methods. Please note high ranking of TLR genes (*TLR5*, *TLR8*, *TLR3* and *TLR7*) in the pathway of Response to Mechanical Stimulus, *CFTR* gene in Recognition for Clathrin-mediated endocytosis, *RNASEL*, *TYK2*, *OAS1* and *OAS3* genes in Interferon alpha-beta signaling. Note also the presence of the relevant pathway of Exhaust vs Memory CD8 T cell Up that also includes *TLR7* gene.

### COVID-19 post-Mendelian model predictivity

The functional interpretation of the variants identified by the feature selection approach, complemented by the strong link between the involved human biological pathways and COVID-19 pathogenicity, support the hypothesis that the *IPGS* equation developed here may contribute significantly to predicting the severity of COVID-19. This hypothesis was tested by using a logistic regression model that predicts COVID-19 severity based on age, sex and the the *IPGS* (after percentile normalization). The training set is composed of 466 patients not included in the training set previously exploited for the *IPGS* feature engineering. The model’s performance was then tested using three independent cohort sets of European ancestry (**Fig. 5A**). The model exhibited an overall accuracy of 0.73, precision equal to 0.78, with a sensitivity and specificity of 0.72 and 0.75, respectively. Noteworthy, all the aforementioned metrics are higher than the corresponding values obtained using a logistic model that adopted as input features only age and sex. The increase in performances of the model with *IPGS* suggests that this score indeed confers significant additional (genetic) information for predicting COVID-19 severity compared to only age and sex. The increase of the performances is statistically significant (p-value < 0.05 for accuracy, precision, sensitivity, specificity) with respect to the distribution of performances for an ensemble of models where the *IPGS* feature has been randomized (**Fig. 5C** **lower left**). A third logistic regression model fitted with *IPGS* alone, shows performances well above the random guess. Furthermore, the empirical probability density function of *IPGS* scores (**Fig. 5C** **right**) has been estimated for the severe and non-severe patients of the cohort including both training and testing sets. It is worth noting the shift on the right of the *IPGS* distribution for the severe patients, with significant p-value (<0.001) for the t-test of mean difference. This difference between severe and non-severe cases is preserved for the male and female cohorts when analyzed separately (p-values <0.001 and 0.024, respectively).

**Figure 5.**
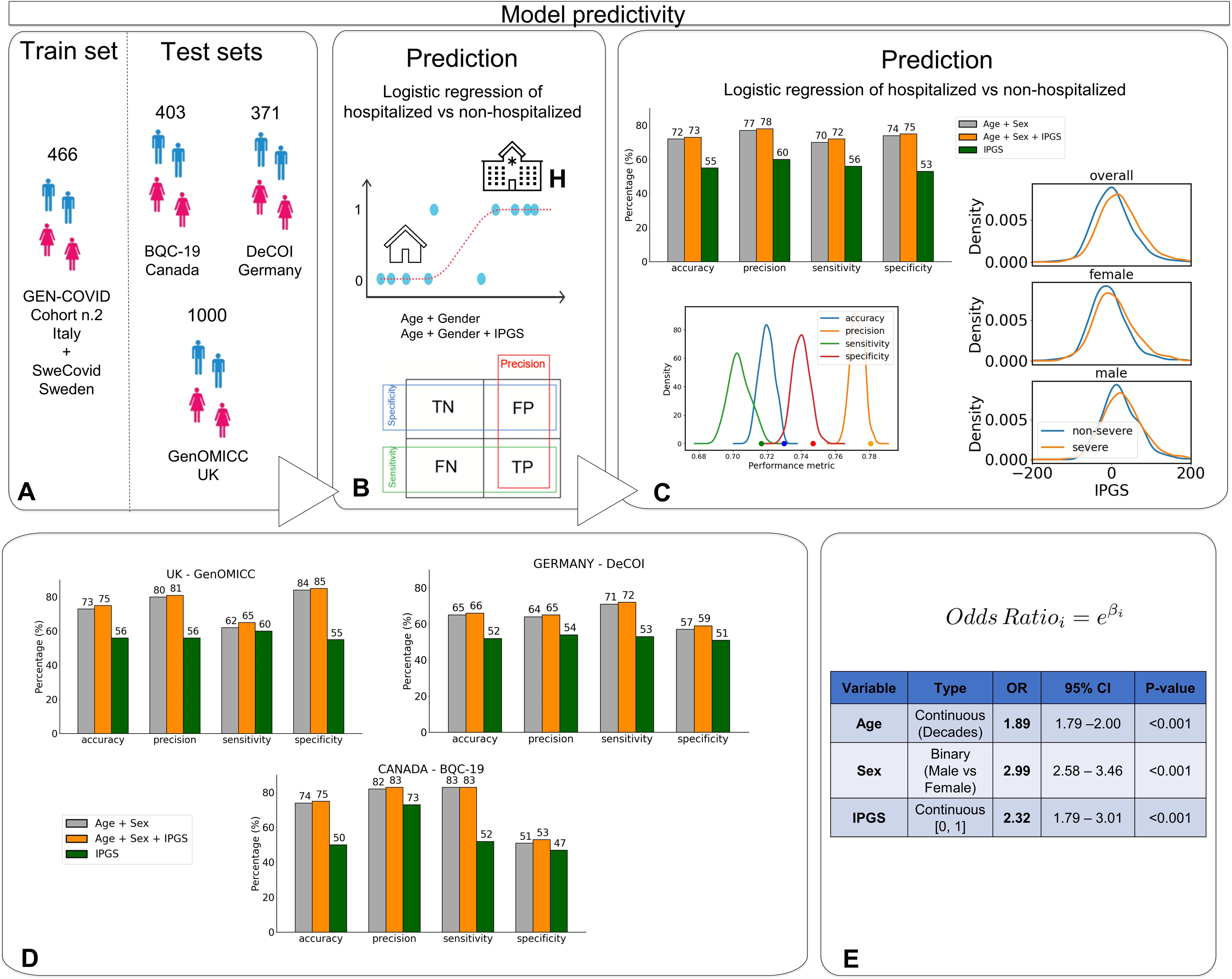
Model predictivity. **A)** The post-Mendelian model was trained using a sample of 466 patients from the GEN-COVID cohort n.2 and Swedish cohort (having cases only) and tested with three additional European cohorts from UK, Germany and Canada. **B)** A logistic regression model was used for severity prediction. Severity was defined mainly on the basis of hospitalisation versus not hospitalisation. Hospitalised cases without respiratory support were included in controls. TN=True Negative; TP=True Positive; FN=False Negative; FP=False Positive. **C)** When the IPGS is added to age and gender as a regressor, the performances of the model increase: accuracy +1%, precision +1%, sensitivity +2%, specificity +1%. These increases are statistically significant (p-value <0.05 for accuracy, precision, sensitivity and specificity) with respect to the null distribution obtained by randomizing the *IPGS*. The performances of the model built with IPGS alone are all above the random guess. In addition, on the right we reported the distributions of the *IPGS* for severe and non-severe patients. **D)** In the three tested cohorts, when the *IPGS* is added to age and sex as a regressor, all the performances increase: the accuracy up to +2%, the precision up to +1%, the sensitivity up to +3%, and the specificity up to +2%. We conclude that *IPGS* is able to improve prediction of clinical outcome in addition to the well-established powerful factors of age and sex. **E)** The univariate logistic regression models fitted on the cohort including both train and test, confirmed that the *IPGS* is associated with severity with an odds-ratio (OR) of 2.32, while age (continuous in decades) and sex have an OR of 1.89 and 2.99, respectively.

In line with the results obtained using the overall test set, the model including *IPGS*, age, and sex performed better than the model considering only sex and age as inputs, in each of the testing cohorts, separately (**Fig. 5D**). The increase in performance was systematically observed throughout all the cohorts: on average +1.33% for accuracy, +1% for precision, +1.33% for sensitivity, +1.67% for specificity. Considering the difference in phenotype classification inherent to a comparison among various international cohorts, and the genetic variability among different European sub-populations, the consistent increase in performances observed for the model with *IPGS* demonstrates that this score provides a robust index for predicting COVID-19 severity. As a further test for the importance of the *IPGS* score for predicting COVID-19 severity, the univariate logistic models were used on the overall set including both train and test cohorts to estimate the OR of severe COVID-19 for *IPGS*, age, and sex, separately. The test confirmed that severity was associated with *IPGS,* showing an OR of 2.32 (p<0.001, 95% confidence interval [1.79, 3.01]) with age, measured in decades, and sex, having OR of 1.89 (p<0.001, 95% confidence interval [1.79, 2.00]) and 2.99 (p<0.001, 95% confidence interval [2.58, 3.46]) respectively (**Fig. 5E**). The multivariate logistic regression using sex, age, and *IPGS* together, provided similar results reported in **Supplementary Table 9** confirming the goodness of the regressors’ OR. When adjusting for comorbidities, in the train cohort where the comorbidities were available, with a multivariable logistic model, OR of IPGS was 2.46 (p=0.05, 95% confidence interval [1.15, 5.25]) as shown in **Supplementary Table 10**. This result further confirms that IPGS is a reliable predictor of COVID-19 clinical severity.

### Advantages of IPGS and clinical interpretability of connected features

We then wanted to compare the clinical outcome with the probability of severity obtained from three different models: IPGS alone, sex-age alone or combined model (represented as heatmap in **Fig. 6**). It appears evident that in a subset of patients, the 2 models based on sex-age alone and IPGS alone have a discordant prediction (left and right end of dendrogram in **Fig. 6A**). In these cases IPGS appears to be a relevant predictor of severity (**Fig. 6A**). This is in accordance with the above presented logistic regression analysis (**Fig. 5E**) that shows IPGS having an OR of 2.32 for severity. Moreover, the list of features on which the IPGS score is built, represent a biological handle for pathophysiological mechanisms and possible personalized adjuvant treatments.

**Figure 6.**
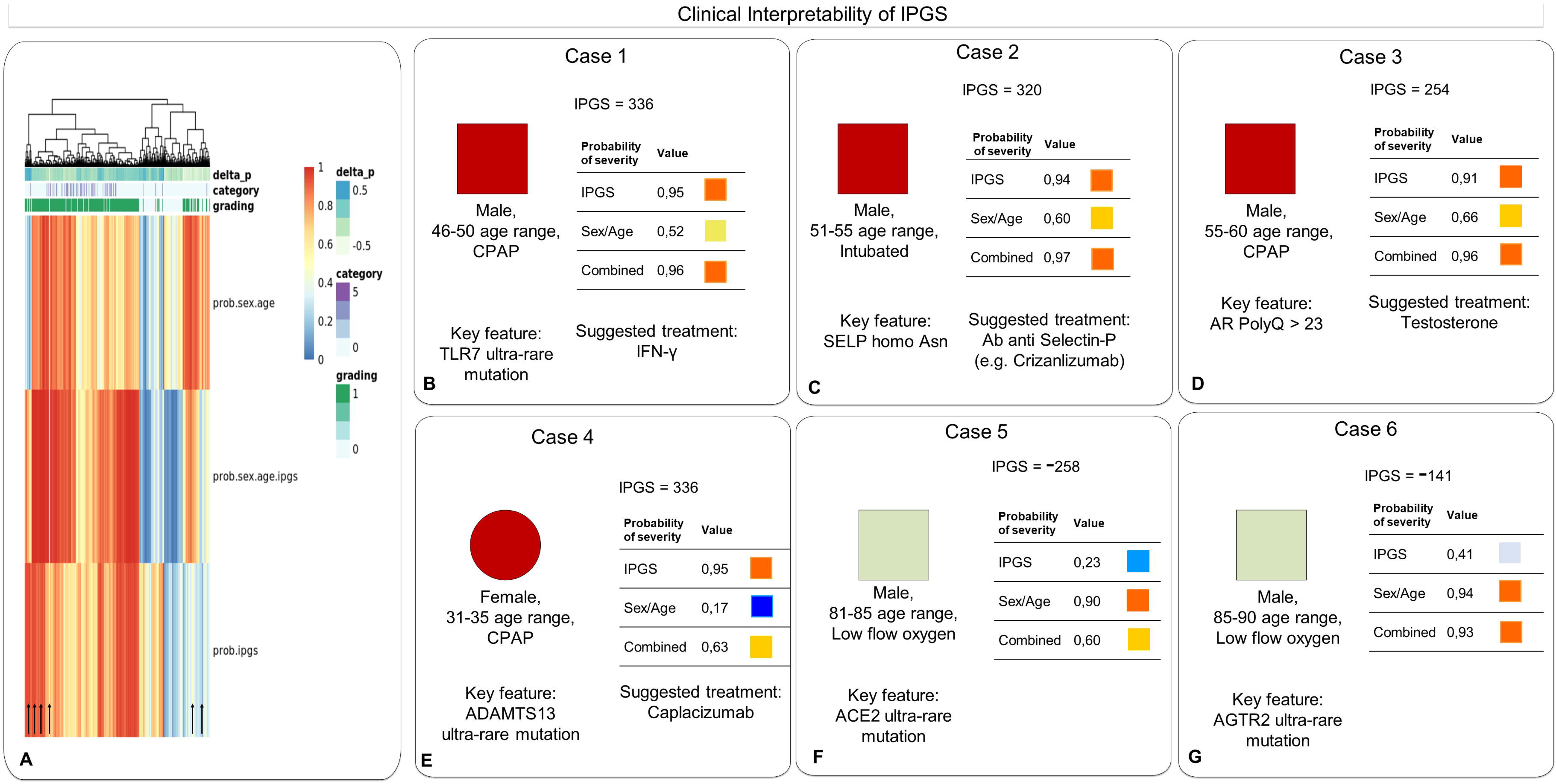
Clinically interpretability of IPGS. Panel **A** shows the GEN-COVID cohort dendrogram and heatmaps of the probabilities of severity based on the 3 different models: sex-age alone, IPGS alone and combined model. In the extreme ends of dendrogram (left and right) the probability of severity based on sex-age alone and IPGS alone is highly discordant (different colours). Selected examples corresponding to the arrows are illustrated in panels B-G. In each panel IPGS score, probabilities of severity and key features useful for bedside clinical management are shown. **B**) Male patient, in the 46-50 age range, treated with CPAP ventilation, tocilizumab, enoxaparin, hydroxychloroquine and lopinavir/ritonavir; no comorbidities except for asthma have been reported. The patient presented a rare *TLR7* mutation that leads to an impaired production of interferon gamma [12]. **C**) Male patient, in the 51-55 age range, treated with invasive mechanical ventilation, steroids and enoxaparin. He had among comorbidities obesity, anxiety, hypertension and cerebral ischemia. He was found to be homozygous for the *SELP* rs6127 (p.Asp603Asn). Homozygosity of Asparagine in position 603 of Selectin P makes this endothelial proteine more prone to clot formation and male patients more prone to COVID-19 thrombosis [18]. Hence the rationale for considering as putative adjuvant therapy in the management of similar cases the anti Selectin P antibodies, a drug already approved for vascular events of sickle cell anemia. **D**) Male patient, in the 51-55 age range, treated with CPAP ventilation, tocilizumab, steroids, enoxaparin, hydroxychloroquine and lopinavir/ritonavir; no comorbidities except for diabetes. He was found to have the androgen receptor polyQ repeats ≥23. The regular function of the androgen receptor is correlated with a beneficial immunomodulatory effect in those male patients in whom the increase in testosterone levels may overcome the receptor resistance. The rationale is to consider giving testosterone to those male subjects who cannot, on their own, raise the levels enough to overcome the receptor resistance due to poly-glutamine stretch longer than 23 repeats [16]. **E**) Female patient, in the 31-35 age range, treated with CPAP ventilation and steroids, enoxaparin and azithromycin; no comorbidities except for hypothyroidism. She was a carrier of an ultra rare mutation in *ADAMTS13*. Impaired function of ADAMTS13 leads to reduced cleavage of von Willebrand factor (vWF) and enhanced clot formation. The effect is enhanced in females and responsible for SARS-CoV-2 related thrombosis. Anti vWF immunoglobulines would be a putative therapeutic option to consider in similar cases. **F-G**) examples of low IPGS and related key features. **F)** Male patient, in the 81-85 age range, treated with low-flow oxygen. No information regarding pharmacological therapy during hospitalization is present. Among comorbidities: diabetes mellitus, congestive heart failure and bowel cancer and steroids. He presented an ultra rare mutation in *ACE2*. **G)** Male patient, in the 86-90 age range, treated with low-flow oxygen, steroid, enoxaparin and ceftriaxone plus azithromycin. Among his comorbidities: colon diverticulosis with constipation?, benign prostatic hyperplasia?, anxious-depressive syndrome, sideropenic anemia. He was a carrier of an ultra rare mutation in *AGTR2*.

For example, three male patients, within two distinct age ranges (46-50, 51-55) (**panels B, C and D)** with severe outcome (intubation and CPAP) are imperfectly represented by the sex-age model (probability of severity from 0,52 to 0,66) and better represented by the IPGS model (probability of severity from 0,91 to 0,95). The detected genetic variants that would allow to clinically consider putative personalized treatments in similar cases are: i) *TLR7* ultra rare mutation indicating to consider possible adjuvant treatment with IFN gamma administration [12]; ii) homozygosity 603Asn in *SELP* gene suggesting putative adjuvant treatment with anti selectin P autoantibodies (e.g. Crizanlizumab)[18] and iii) polyQ longer than 23 in *AR* gene suggesting to consider possible adjuvant treatment with testosterone [16].

In a female patient, within age range 31-35, the sex-age model showed a probability of severity of 0.17 (**panel D**) while the IPGS score was 336 corresponding to a probability of severity of 0.95. The patient had no comorbidities except for hypothyroidism. She underwent steroid treatment and CPAP ventilation. She was found to be carrier of *ADAMTS13* ultra rare mutation, being more susceptible to thrombosis (due to reduced capacity of cutting von Willebrand factor); she had indeed a high D-dimer value. Caplacizumab (an antibody anti vWF) would be an option to consider as possible adjuvant treatment in the clinical management of similar cases

Two male patients, within two distinct age ranges (81-85, 86-90) (**panel F and G**) with a relatively mild respiratory outcome (hospitalised with low flow oxygen therapy) presented an IPGS score of -258 and -141 respectively. Their severity probabilities calculated on sex-age (0.9 and 0.94) do not mirror the relatively mild clinical outcome, which is instead better represented by the severity probability calculated in IPGS only (0.23 and 0.41). Those two patients presented ultra-rare variants in *ACE2* gene, likely responsible for reduced viral load [35], and in *AGTR2* gene, which reduced activity is known to prevent cystic fibrosis pulmonary manifestation [36].

## Discussion

The importance of combination of rare and low frequency variants has already been demonstrated to contribute to the prediction of susceptibility in other complex disorders [37–38]. Here we further expand this approach while demonstrating that ultra-rare, rare, low frequency, as well as common variants contribute to the likelihood of developing a severe form of COVID-19. Furthermore, we included in our analyses a calibration of the relative weight of the variants *vis-a-vis* their impact on disease severity: a single ultra-rare variant might well by itself cause a severe phenotype of COVID-19, while this is less probable for a common polymorphism, one that is likely to have a markedly less direct effect on protein functionality. We performed a first modellization of COVID-19 genetics using both rare and common variants [20]. Because feature selection methodologies are generally sensitive to allele frequency, the extraction was performed separately for rare (MAF <1/100) and common (MAF >1/100) variants. However, the methodology revealed the insight that low-frequency variants (MAF from 1% to 5%) were disadvantaged if selected together with common ones. Furthermore, for extracting Mendelian-like genes a threshold of MAF < 0.1% (ultra-rare variants) appeared more effective than MAF< 1% and all mutations in the *TLR7* gene that proved to have loss of function had indeed MAF< 1/1000 [12]. The model we arrived at, now considers separately ultra-rare, rare, low-frequency, and common variants.

Similar to the classical PRS (Polygenic Risk Score), the proposed IPGS (*Integrated* PolyGenic Score) may prove reliable for assessing the probability of severe COVID-19 following infection by SARS-CoV-2 [21]. While PRS is based on common polymorphisms found at the genomic level with the majority of loci potentially conferring risk being not easily interpretable due to the uncertainty of linked genes, IPGS allows immediate biological interpretation because it only includes coding variants. Furthermore, as opposed to PRS, IPGS relies on both polymorphisms and rare variants is capable of differentially weighting features in an indirectly proportional way in respect to frequency and therefore to protein impact. Each patient indeed is assigned both a number and the list of her/his common and low-frequency polymorphisms relevant to COVID-19 supported by medically actionable information and of rare and ultra-rare variants conferring either risk of severity or protection from severe disease. Drawing on the entire picture presented through IPGS analysis, personalized adjuvant therapy could be envisaged. At the time of writing, a platform trial based on genetic markers is being discussed with the Italian Medicines Agency (EudraCT Number: 2021-002817-32).

Within 25 reported genomic SNPs demonstrably related to COVID-19 susceptibility/severity, 5 were reported to be in LD with coding variants [9, 39]. The model presented here might provide useful information for uncovering the identity of the gene/coding variants responsible for COVID-19 susceptibility/severity linked to these genomic SNPs (**Fig. 2D**). For example, on chromosome 12, the genes mapping to the locus tagged by rs10774671 [9] are both *OAS1* and *OAS3*. In *OAS3* the coding variant is an Arginine to Lysine substitution (rs1859330) in high LD (0.8) with the tag SNP. This polymorphism was already associated with viral infection [40] based on the presence of Lysine having been shown to lead to a decreased INF-γ production. In *OAS1* the haplotype (including 4 missense variants: G162S, A352T, R361T, and G397R), the splicing variant 1039-G>A (the reported genomic polymorphism itself), and the truncating mutation T359fs*26 are associated with severity and predicted to impair OAS1 function. Both OAS1 and OAS3 induce RNASEL, which in turn exerts antiviral activity. Further support for the role of the OAS/RNASEL axis is indicated by the presence of ultra-rare recessive variants.

This innovative approach allowed us to better select genes located on the X chromosome related to COVID-19 that affect males and females in opposite ways (**Fig. 2A** and **Supplementary Tables 3 and 6**). Interestingly, many of these genes were previously confirmed or hypothesized to escape the X chromosome inactivation. With respect to these genes, females produce twice the levels of protein in comparison with males. Mutations in hemizygous state in males and heterozygous state in females appear silent until SARS-CoV-2 infection occurs. For example, *TLR7* and *TLR8* are selected for ultra-rare and associated with severity in males and with protection from severe disease in heterozygous females. We know that the activation of *TLR7/8* induces the production of type 1 and type 2 IFN as well as pro-inflammatory cytokines, where the production defect in hemizygous males leads to severe COVID-19. However, an excess of the sensor can also lead to damage from hyperinflammation. Therefore, the condition of carrier females is the more favorable state and has in fact been associated with mild COVID-19 [41] .

Pathway analysis pointed to the relevance of obvious actors in COVID-19 pathology, such as immune cells and interferon signaling, but also to the important role of specific organs (brain, digestive tract, kidney, reproductive system) and functions (metabolism of lipids and steroids). The pathways identified through GSEA analyses reflected the multi-organ nature of the disease. In addition, our analyses reveal new candidate determinants of disease variability. The four pathways linked to cilium motility suggest a role for ciliated cells of the respiratory tract (and possibly others) in antiviral defense. The functionality of the clathrin-mediated endocytosis pathway may likely affect viral entry [42]. Likewise, endoplasmic reticulum associated protein degradation (ERAD), which is linked to autophagy and SARS-CoV-2 life-cycle [43], may also be relevant. Other pathways with a less obvious but potentially interesting role in the disease include cell adhesion and mechanical stimulus signaling.

The strong link between the involved human biological pathways and COVID-19 pathogenicity support the hypothesis that the proposed *IPGS* equation may contribute significantly to predicting the disease severity of COVID-19. Indeed, an overall significant increase of performance was obtained in comparison with the model based on solely on age and sex. Furthermore, the *IPGS* is significantly associated with severity, showing an OR of 2.46 after adjustment for age, sex, and comorbidities. This indicates that IPGS is a novel prognostic factor that should be considered in the management of COVID-19 patients.

Modelling precisely the role of the entire range of host genomics affecting disease susceptibility and severity in COVID-19 is critical to obtaining a complete biological understanding of the aetiology and pathogenicity of COVID-19 as well as other severe complex diseases. The application of *IPGS* based on Machine Learning principles within a post-Mendelian model allows us to more precisely identify the gene variants at play in COVID-19 as well as their specific roles, individually and in combination. This deep dive into the genetic architecture that allows for, contributes to, or even helps prevent diseases while increasing or decreasing their impact is critical for, and directly translatable into, (personalized) medicines development as well as prevention and treatment protocols. An integrated modelling of genetic variants based on a limited patient cohort, even limited in its geographical spread, may be sufficient for the development of diagnosis, and therapeutics across a wider range of populations. The advantage of this *IPGS* post-Mendelian model is that it learns and continues to learn as well as being a model from which we can obtain insights on the fundamental architecture of human genomics when confronted with severe and complex diseases.

## Data Availability

The data and samples referenced here are housed in the GEN-COVID Patient Registry and the GEN-COVID Biobank and are available for consultation. You may contact the corresponding author, Prof. Alessandra Renieri.

## ACKNOWLEDGMENTS

This study is part of the GEN-COVID Multicenter Study, https://sites.google.com/dbm.unisi.it/gen-covid, the Italian multicenter study aimed at identifying the COVID-19 host genetic bases. Specimens were provided by the COVID-19 Biobank of Siena, which is part of the Genetic Biobank of Siena, member of BBMRI-IT, of Telethon Network of Getic Biobanks (project no. GTB18001), of EuroBioBank and of D-Connect. We thank the CINECA consortium for providing computational resources and the Network for Italian Genomes (NIG) http://www.nig.cineca.it for its support. We thank private donors for the support provided to A.R. (Department of Medical Biotechnologies, University of Siena) for the COVID-19 host genetics research project (D.L n.18 of March 17, 2020). We also thank the COVID-19 Host Genetics Initiative (https://www.covid19hg.org/). We thank Uppsala Intensive Care COVID-19 research group; Tomas Luther, Anders Larsson, Katja Hanslin, Anna Gradin, Sarah Galien, Sara Bulow Anderberg, Jacob Rosén and Sten Rubertsson. We thank the patients and their loved ones who volunteered to contribute to this study at one of the most difficult times in their lives, and the research staff in every intensive care unit who recruited patients at personal risk during the most extreme conditions we have ever witnessed in UK hospitals. Whole-genome sequencing was done by Illumina in partnership with the University of Edinburgh and Genomics England and was funded by UK Department of Health and Social Care, UKRI and LifeArc. The views expressed are those of the authors and not necessarily those of the DHSC, DID, NIHR, MRC, Wellcome Trust or PHE. The Health Research Board of Ireland (Clinical Trial Network Award 2014-12) funds collection of samples in Ireland. This study owes a great deal to the National Institute of Healthcare Research Clinical Research Network (NIHR CRN) and the Chief Scientist Office (Scotland), who facilitate recruitment into research studies in NHS hospitals, and to the global ISARIC and InFACT consortia

## Supplementary Materials

**Supplementary Figure 1. Barplots for 0.01 both**

Barplot of significance values (NOM p-values, -log_10_ transformation) from GSEA analysis for all the pathways significant in both females (orange) and males (blue), p<0.01. Vertical dotted line indicates the adopted significance threshold.

**Supplementary Figure 2. Representative heatmaps for 0.01_both**

Heatmaps of the genes belonging to representative pathways significant in both females and males, p<0.01. The color gradient represents the weight of each gene, calculated as described in methods.

**Supplementary Figure 3. Barplots for 0.005_any**

Barplot of significance values (NOM p values, -log_10_ transformation) from GSEA analysis for all the pathways significant in either females (orange) or males (blue), p<0.005. Vertical dotted line indicates the adopted significance threshold.

**Supplementary Figure 4. Representative heatmaps for 0.005_any**

Heatmaps of the genes belonging to representative pathways significant in either females or males, p<0.005. The color gradient represents the weight of each gene, calculated as described in methods.

**Supplementary Table 1.** Cohorts demography: information regarding studies contributing to this study.

**Supplementary Table 2.** Post-Mendelian model: boolean representations

**Supplementary Table 3a-b.** Ultra-rare features extracted in males; Ultra-rare features extracted in females

**Supplementary Table 4a-b.** Rare features extracted in males; rare features extracted in females

**Supplementary Table 5a-b.** Low-frequency features extracted in males; low-frequency features extracted in females

**Supplementary Table 6a-b.** Common features extracted in males; Common features extracted in females

**Supplementary Table 7.** List of genes and weights used for pathway analysis

**Supplementary Table 8.** List of genes belonging to the significant pathways and their weights in Females and Males

**Supplementary Table 9.** Association study with multivariate logistic regression

**Supplementary Table 10.** Association study with multivariate logistic regression and comorbidities

**Supplementary Table 11.** Common coding variants in LD with previously reported genomic SNPs

## AUTHOR CONTRIBUTIONS

Conceptualization, Francesca Mari, Maddalena Fratelli, Simone Furini and Alessandra Renieri; Data curation, Ilaria Meloni, Susanna Croci, Maria Palmieri, Mirjam Lista, Giada Beligni; Formal analysis, Nicola Picchiotti, Elisa Benetti, Chiara Fallerini, Sergio Daga, Floriana Valentino, Margherita Baldassarri, Francesca Fava, Kristina Zguro, Marco Tanfoni, Francesca Colombo, Enrico Cabri, Maddalena Fratelli, Chiara Gabbi, Elisa Frullanti, GEN-COVID Multicenter Study, Marco Gori, Alessandra Renieri and Simone Furini; Funding acquisition, Alessandra Renieri and Francesca Mari; Methodology, Nicola Picchiotti, Elisa Benetti, Marco Tanfoni, Francesca Colombo, Enrico Cabri, Maddalena Fratelli, Chiara Gabbi, Stefania Mantovani, Sara Amitrano, Mirella Bruttini and Simone Furini; Project administration, Francesca Mari and Alessandra Renieri; Supervision, Francesca Mari, GEN-COVID Multicenter Study and Alessandra Renieri; Validation, Simone Furini, Guillaume Butler-Laporte, Brent Richards, Hugo Zeberg, Miklos Lipcsey, Michael Hultstrom, Kerstin U. Ludwig, Eva C. Schulte, Erola Pairo-Castineira, Mark Caulfield, Loukas Moutsianas, Athanasios Kousathanas, Susan Walker, John Kenneth Baillie, Axel Schmidt, Robert Frithiof; Writing – original draft, Nicola Picchiotti, Elisa Benetti, Chiara Fallerini, Sergio Daga, Margherita Baldassarri, Francesca Fava, Kristina Zguro, Sara Amitrano, Mirella Bruttini, Maria Palmieri, Mirjam Lista, Giada Beligni, Floriana Valentino, Susanna Croci, Ilaria Meloni, Francis P. Crawley, Marco Tanfoni, Francesca Colombo, Enrico Cabri, Maddalena Fratelli, Chiara Gabbi, Elisa Frullanti, Francesca Mari, GEN-COVID Multicenter Study, Marco Gori, Alessandra Renieri and Simone Furini.

## INSTITUTIONAL REVIEW BOARD STATEMENT

The study was conducted according to the guidelines of the Declaration of Helsinki. The GEN-COVID is a multicentre academic observational study that was approved by the Internal Review Boards (IRB) of each participating centre (protocol code 16917, dated March 16, 2020 for GEN-COVID at the University Hospital of Siena). BQC19 received ethical approval from the Jewish General Hospital (JGH) research ethics board (2020-2137). The Swedish part of the study was performed at the general intensive care unit (ICU) of Uppsala University Hospital, Sweden (a tertiary hospital) and approved by the National Ethical Review Agency (No. 2020-01623). The Declaration of Helsinki and its subsequent revisions were followed. DeCOI received ethical approval by the Ethical Review Board (ERB) of the participating hospitals/centres (Technical University Munich, Munich, Germany; Medical Faculty Bonn, Bonn, Germany; Medical Board of the Saarland, Germany; University Duisburg-Essen, Germany; Medical Faculty Duesseldorf, Düsseldorf, Germany). GenOMICC and ISARIC4C were approved by the appropriate research ethics committees (Scotland, 15/SS/0110; England, Wales and Northern Ireland, 19/WM/0247).

## INFORMED CONSENT STATEMENT

The patients were informed of this research and agreed to it through the informed consent process.

## DATA AVAILABILITY AND DATA SHARING STATEMENT

**❖ GEN-COVID Multicenter Study (https://sites.google.com/dbm.unisi.it/gen-covid)**

Francesca Montagnani^1,27^, Mario Tumbarello^1,27^, Ilaria Rancan^1,27^, Massimiliano Fabbiani^27^, Barbara Rossetti^27^, Laura Bergantini^28^, Miriana D’Alessandro^28^, Paolo Cameli^28^, David Bennett^28^, Federico Anedda^29^, Simona Marcantonio^29^, Sabino Scolletta^29^, Federico Franchi^29^, Maria Antonietta Mazzei^30^, Susanna Guerrini^30^, Edoardo Conticini^31^, Luca Cantarini^31^, Bruno Frediani^31^, Danilo Tacconi^32^, Chiara Spertilli Raffaelli^32^, Marco Feri^33^, Alice Donati^33^, Raffaele Scala^34^, Luca Guidelli^34^, Genni Spargi^35^, Marta Corridi^35^, Cesira Nencioni^36^, Leonardo Croci^36^, Gian Piero Caldarelli^37^, Maurizio Spagnesi^38^, Davide Romani^38^, Paolo Piacentini^38^, Maria Bandini^38^, Elena Desanctis^38^, Silvia Cappelli^38^, Anna Canaccini^39^, Agnese Verzuri^39^, Valentina Anemoli^39^, Manola Pisani^39^, Agostino Ognibene^40^, Alessandro Pancrazzi^40^, Maria Lorubbio^40^, Massimo Vaghi^41^, Antonella D’Arminio Monforte^42^, Federica Gaia Miraglia^42^, Mario U. Mondelli^43,44^,, Massimo Girardis^45^, Sophie Venturelli^45^, Stefano Busani^45^, Andrea Cossarizza^46^, Andrea Antinori^47^, Alessandra Vergori^47^, Arianna Emiliozzi^47^, Stefano Rusconi^48,49^, Matteo Siano^49^, Arianna Gabrieli^49^, Agostino Riva^48,49^, Daniela Francisci^50,51^, Elisabetta Schiaroli^50^, Francesco Paciosi^50^, Andrea Tommasi^50^, Pier Giorgio Scotton^52^, Francesca Andretta^52^, Sandro Panese^53^, Stefano Baratti^53^, Renzo Scaggiante^54^, Francesca Gatti^54^, Saverio Giuseppe Parisi^55^, Francesco Castelli^56^, Eugenia Quiros-Roldan^56^, Melania Degli Antoni^56^, Isabella Zanella^57,58^, Matteo Della Monica^59^, Carmelo Piscopo^59^, Mario Capasso^60,61,62^, Roberta Russo^60,61^, Immacolata Andolfo^60,61^, Achille Iolascon^60,61, 61^, Giuseppe Fiorentino^63^, Massimo Carella^64^, Marco Castori^64^, Filippo Aucella^65^, Pamela Raggi^66^, Rita Perna^66^, Matteo Bassetti^67,68^, Antonio Di Biagio^67,68^, Maurizio Sanguinetti^69,70^, Luca Masucci^69,70^, Alessandra Guarnaccia^69^, Serafina Valente^71^, Oreste De Vivo^71^, Gabriella Doddato^1,2^, Rossella Tita^5^, Annarita Giliberti^1,2^, Maria Antonietta Mencarelli^5^, Caterina Lo Rizzo^5^, Anna Maria Pinto^5^, Valentina Perticaroli^1,2,5^, Francesca Ariani^1,2,5^, Miriam Lucia Carriero^1,2^, Laura Di Sarno^1,2^, Diana Alaverdian^1,2^, Elena Bargagli^28^, Marco Mandalà^72^, Alessia Giorli^72^, Lorenzo Salerni^72^, Patrizia Zucchi^73^, Pierpaolo Parravicini^73^, Elisabetta Menatti^74^, Tullio Trotta^75^, Ferdinando Giannattasio^75^, Gabriella Coiro^75^, Fabio Lena^76^, Gianluca Lacerenza^76^, Domenico A. Coviello^77^, Cristina Mussini^78^, Enrico Martinelli^79^, Sandro Mancarella^80^, Luisa Tavecchia^80^, Mary Ann Belli^80^, Lia Crotti^81,82,83,84,85^, Gianfranco Parati^81,67^, Maurizio Sanarico^86^, Francesco Raimondi^87^, Filippo Biscarini^88^, Alessandra Stella^88^, Marco Rizzi^89^, Franco Maggiolo^89^, Diego Ripamonti^89^, Claudia Suardi^90^, Tiziana Bachetti^91^, Maria Teresa La Rovere^92^, Simona Sarzi-Braga^93^, Maurizio Bussotti^94^, Katia Capitani^3,95^, Simona Dei^96^, Sabrina Ravaglia^97^, Rosangela Artuso^98^, Elena Andreucci^98^, Giulia Gori^98^, Angelica Pagliazzi^98^, Erika Fiorentini^98^, Antonio Perrella^99^, Francesco Bianchi^1,99^, Paola Bergomi^100^, Emanuele Catena^100^, Riccardo Colombo^100^, Sauro Luchi^101^, Giovanna Morelli^101^, Paola Petrocelli^101^, Sarah Iacopini^101^, Sara Modica^101^, Silvia Baroni^102^, Francesco Vladimiro Segala^103^, Francesco Menichetti^104^, Marco Falcone^104^, Giusy Tiseo^104^, Chiara Barbieri^104^, Tommaso Matucci^104^, Davide Grassi^105^, Claudio Ferri^106^, Franco Marinangeli^107^, Francesco Brancati^108^, Antonella Vincenti^109^, Valentina Borgo^109^, Lombardi Stefania^109^, Mirco Lenzi^109^, Massimo Antonio Di Pietro^110^, Francesca Vichi^110^, Benedetta Romanin^110^, Letizia Attala^110^, Cecilia Costa^110^, Andrea Gabbuti^110^, Menè Roberto^81,82^, Umberto Zuccon^111^, Lucia Vietri^111^, Stefano Ceri^112^, Pietro Pinoli^112^, Patrizia Casprini^113^, Giuseppe Merla^114,115^, Gabriella Maria Squeo^114^, Marcello Maffezzoni^116^, Raffaele Bruno^117,118^, Marco Vecchia^117,^, Marta Colaneri^117^, Serena Ludovisi^119^

27) Department of Medical Sciences, Infectious and Tropical Diseases Unit, Azienda Ospedaliera Universitaria Senese, Siena, Italy

28) Unit of Respiratory Diseases and Lung Transplantation, Department of Internal and Specialist Medicine, University of Siena, Italy

29) Dept of Emergency and Urgency, Medicine, Surgery and Neurosciences, Unit of Intensive Care Medicine, Siena University Hospital, Italy

30) Department of Medical, Surgical and Neuro Sciences and Radiological Sciences, Unit of Diagnostic Imaging, University of Siena, Italy

31) Rheumatology Unit, Department of Medicine, Surgery and Neurosciences, University of Siena, Policlinico Le Scotte, Italy

32) Department of Specialized and Internal Medicine, Infectious Diseases Unit, San Donato Hospital Arezzo, Italy

33) Dept of Emergency, Anesthesia Unit, San Donato Hospital, Arezzo, Italy

34) Department of Specialized and Internal Medicine, Pneumology Unit and UTIP, San Donato Hospital, Arezzo, Italy

35) Department of Emergency, Anesthesia Unit, Misericordia Hospital, Grosseto, Italy

36) Department of Specialized and Internal Medicine, Infectious Diseases Unit, Misericordia Hospital, Grosseto, Italy

37) Clinical Chemical Analysis Laboratory, Misericordia Hospital, Grosseto, Italy

38) Dipartimento di Prevenzione, Azienda USL Toscana Sud Est, Italy

39) Dipartimento Tecnico-Scientifico Territoriale, Azienda USL Toscana Sud Est, Italy

40) Clinical Chemical Analysis Laboratory, San Donato Hospital, Arezzo, Italy

41) Chirurgia Vascolare, Ospedale Maggiore di Crema, Italy

42) Department of Health Sciences, Clinic of Infectious Diseases, ASST Santi Paolo e Carlo, University of Milan, Italy

43) Division of Infectious Diseases and Immunology, Department of Medical Sciences and Infectious Diseases, Fondazione IRCCS Policlinico San Matteo,Pavia, Italy

44) Department of Internal Medicine and Therapeutics, University of Pavia, Italy

45) Department of Anesthesia and Intensive Care, University of Modena and Reggio Emilia, Modena, Italy

46) Department of Medical and Surgical Sciences for Children and Adults, University of Modena and Reggio Emilia, Modena, Italy

47) HIV/AIDS Department, National Institute for Infectious Diseases, IRCCS, Lazzaro Spallanzani, Rome, Italy

48) III Infectious Diseases Unit, ASST-FBF-Sacco, Milan, Italy

49) Department of Biomedical and Clinical Sciences Luigi Sacco, University of Milan, Milan, Italy

50) Infectious Diseases Clinic, Department of Medicine 2, Azienda Ospedaliera di Perugia and University of Perugia, Santa Maria Hospital, Perugia, Italy

51) Infectious Diseases Clinic, “Santa Maria” Hospital, University of Perugia, Perugia, Italy

52) Department of Infectious Diseases, Treviso Hospital, Local Health Unit 2 Marca Trevigiana, Treviso, Italy

53) Clinical Infectious Diseases, Mestre Hospital, Venezia, Italy.

54) Infectious Diseases Clinic, ULSS1, Belluno, Italy

55) Department of Molecular Medicine, University of Padova, Italy

56) Department of Infectious and Tropical Diseases, University of Brescia and ASST Spedali Civili Hospital, Brescia, Italy

57) Department of Molecular and Translational Medicine, University of Brescia, Italy;

58) Clinical Chemistry Laboratory, Cytogenetics and Molecular Genetics Section, Diagnostic Department, ASST Spedali Civili di Brescia, Italy

59) Medical Genetics and Laboratory of Medical Genetics Unit, A.O.R.N. “Antonio Cardarelli”, Naples, Italy

60) Department of Molecular Medicine and Medical Biotechnology, University of Naples Federico II, Naples, Italy

61) CEINGE Biotecnologie Avanzate, Naples, Italy

62) IRCCS SDN, Naples, Italy

63) Unit of Respiratory Physiopathology, AORN dei Colli, Monaldi Hospital, Naples, Italy

64) Division of Medical Genetics, Fondazione IRCCS Casa Sollievo della Sofferenza Hospital, San Giovanni Rotondo, Italy

65) Department of Medical Sciences, Fondazione IRCCS Casa Sollievo della Sofferenza Hospital, San Giovanni Rotondo, Italy

66) Clinical Trial Office, Fondazione IRCCS Casa Sollievo della Sofferenza Hospital, San Giovanni Rotondo, Italy

67) Department of Health Sciences, University of Genova, Genova, Italy

68) Infectious Diseases Clinic, Policlinico San Martino Hospital, IRCCS for Cancer Research Genova, Italy

69) Microbiology, Fondazione Policlinico Universitario Agostino Gemelli IRCCS, Catholic University of Medicine, Rome, Italy

70) Department of Laboratory Sciences and Infectious Diseases, Fondazione Policlinico Universitario A. Gemelli IRCCS, Rome, Italy

71) Department of Cardiovascular Diseases, University of Siena, Siena, Italy

72) Otolaryngology Unit, University of Siena, Italy

73) Department of Internal Medicine, ASST Valtellina e Alto Lario, Sondrio, Italy

74) Study Coordinator Oncologia Medica e Ufficio Flussi Sondrio, Italy

75) First Aid Department, Luigi Curto Hospital, Polla, Salerno, Italy

76) Department of Pharmaceutical Medicine, Misericordia Hospital, Grosseto, Italy.

77) U.O.C. Laboratorio di Genetica Umana, IRCCS Istituto G. Gaslini, Genova, Italy

78) Infectious Diseases Clinics, University of Modena and Reggio Emilia, Modena, Italy

79) Department of Respiratory Diseases, Azienda Ospedaliera di Cremona, Cremona, Italy

80) U.O.C. Medicina, ASST Nord Milano, Ospedale Bassini, Cinisello Balsamo (MI), Italy

81) Istituto Auxologico Italiano, IRCCS, Department of Cardiovascular, Neural and Metabolic Sciences, San Luca Hospital, Milan, Italy

82) Department of Medicine and Surgery, University of Milano-Bicocca, Milan, Italy

83) Istituto Auxologico Italiano, IRCCS, Center for Cardiac Arrhythmias of Genetic Origin, Milan, Italy

84) Istituto Auxologico Italiano, IRCCS, Laboratory of Cardiovascular Genetics, Milan, Italy

85) Member of the European Reference Network for Rare, Low Prevalence and Complex Diseases of the Heart-ERN GUARD-Heart

86) Independent Data Scientist, Milan, Italy

87) Scuola Normale Superiore, Pisa, Italy

88) CNR-Consiglio Nazionale delle Ricerche, Istituto di Biologia e Biotecnologia Agraria (IBBA), Milano, Italy

89) Unit of Infectious Diseases, ASST Papa Giovanni XXIII Hospital, Bergamo, Italy

90) Fondazione per la ricerca Ospedale di Bergamo, Bergamo, Italy

91) Direzione Scientifica, Istituti Clinici Scientifici Maugeri IRCCS, Pavia, Italy

92) Istituti Clinici Scientifici Maugeri IRCCS, Department of Cardiology, Institute of Montescano, Pavia, Italy

93) Istituti Clinici Scientifici Maugeri, IRCCS, Department of Cardiac Rehabilitation, Institute of Tradate (VA), Italy

94) Istituti Clinici Scientifici Maugeri IRCCS, Department of Cardiology, Institute of Milan, Milan, Italy

95) Core Research Laboratory, ISPRO, Florence, Italy

96) Health Management, Azienda USL Toscana Sudest, Tuscany, Italy

97) IRCCS C. Mondino Foundation, Pavia, Italy

98) Medical Genetics Unit, Meyer Children’s University Hospital, Florence, Italy

99) Department of Medicine, Pneumology Unit, Misericordia Hospital, Grosseto, Italy.

100) Department of Anesthesia and Intensive Care Unit, ASST Fatebenefratelli Sacco, Luigi Sacco Hospital, Polo Universitario, University of Milan, Milan

101) Infectious Disease Unit, Hospital of Lucca, Italy

102) Department of Diagnostic and Laboratory Medicine, Institute of Biochemistry and Clinical Biochemistry, Fondazione Policlinico Universitario A. Gemelli IRCCS, Catholic University of the Sacred Heart, Rome, Italy.

103) Clinic of Infectious Diseases, Catholic University of the Sacred Heart, Rome, Italy

104) Department of Clinical and Experimental Medicine, Infectious Diseases Unit, University of Pisa, Pisa, Italy

105) Dept. of Clinical Medicine, Public Health, Life and Environment Sciences, University of L’Aquila, L’Aquila, Italy

106) Department of Clinical Medicine, Public Health, Life and Environment Sciences, University of L’Aquila, Italy

107) Anesthesiology and Intensive Care, University of L’Aquila, L’Aquila, Italy

108) Medical Genetics Unit, Department of Life, Health and Environmental Sciences, University of L’Aquila, L’Aquila, Italy

109) Infectious Disease Unit, Hospital of Massa, Italy

110) Infectious Diseases Unit, Santa Maria Annunziata Hospital, USL Centro, Florence, Italy

111) Respiratory Diseases Unit, “Santa Maria degli Angeli” Hospital, Pordenone, Italy

112) Department of Electronics, Information and Bioengineering (DEIB), Politecnico di Milano, Milano, Italy.

113) Laboratory of Clinical Pathology and Immunoallergy, Florence-Prato, Italy

114) Laboratory of Regulatory and Functional Genomics, Fondazione IRCCS Casa Sollievo della Sofferenza, San Giovanni Rotondo (Foggia), Italy

115) Department of Molecular Medicine and Medical Biotechnology, University of Naples Federico II, Naples, Italy.

116) University of Pavia, Pavia, Italy

117) Division of Infectious Diseases I, Fondazione IRCCS Policlinico San Matteo, Pavia, Italy

118) Department of Clinical, Surgical, Diagnostic, and Pediatric Sciences, University of Pavia, Pavia, Italy

119) Fondazione IRCCS Ca’ Granda Ospedale Maggiore Policlinico, Milan, Italy.

**◇ WES/WGS working group within the HGI (https://www.covid19hg.org/projects/)**

**-The genetic predisposition to severe COVID-19 (SweCovid)**

Yanara Marincevic-Zuniga^120^, Jessica Nordlund^120^, Tomas Luther^16^, Anders Larsson^121,^ Katja Hanslin^16^, Anna Gradin^16^, Sarah Galien^16^, Sara Bulow Anderberg^16^, Jacob Rosén^16^, Sten Rubertsson^16^, Hugo Zeberg^15,^, Robert Frithiof^16^, Miklós Lipcsey^16,17^, Michael Hultström^16,18^

120) Department of Medical Sciences, Science for Life Laboratory, Uppsala University, Uppsala, Sweden

121) Department of Medical Sciences, Clinical Chemistry, Uppsala University, Uppsala, Sweden

^❈^ **GenOMICC Consortium**

- **GenOMICC co-investigators**

Sara Clohisey^25^, Peter Horby^122^, Johnny Millar^25^, Julian Knight^123^, Hugh Montgomery^124^, David Maslove^125^, Lowell Ling^126^, Alistair Nichol^127^, Charlotte Summers^128^, Tim Walsh^26^, Charles Hinds^129^, Malcolm G. Semple^130,131^, Peter J.M. Openshaw^132,133^, Manu Shankar-Hari^134^, Antonia Ho^135^, Danny McAuley^1367,1378^, Chris Ponting^24^, Kathy Rowan^1389^, J. Kenneth Baillie^24,25,26^.

- **Central management and laboratory team**

Fiona Griffiths^25^, Wilna Oosthuyzen^25^ Jen Meikle^25^, Paul Finernan^25^, James Furniss^25^, Ellie Mcmaster^25^, Andy Law^25^, Sara Clohisey^25^, J. Kenneth Baillie^24,25,26^, Trevor Paterson^25^, Tony Wackett^25^, Ruth Armstrong^25^, Lee Murphy^139^, Angie Fawkes^139^, Richard Clark^139^, Audrey Coutts^139^, Lorna Donnelly^139^, Tammy Gilchrist^139^, Katarzyna Hafezi^139^, Louise Macgillivray^139^, Alan Maclean^139^, Sarah McCafferty^139^, Kirstie Morrice^139^, Jane Weaver^25^, Ceilia Boz^25^, Ailsa Golightly^25^, Mari Ward^25^, Hanning Mal^25^, Helen Szoor-McElhinney^25^, Adam Brown^25^, Ross Hendry^25^, Andrew Stenhouse^25^, Louise Cullum^25^, Dawn Law^25^, Sarah Law^25^, Rachel Law^25^, Max Head Fourman^25^, Maaike Swets^25^, Nicky Day^25^, Filip Taneski^25^, Esther Duncan^25^, Marie Zechner^25^, Nicholas Parkinson^25^.

**- Data analysis team**

Erola Pairo-Castineira^24,25^, Sara Clohisey^25^, Lucija Klaric^24^, Andrew D. Bretherick^24^, Konrad Rawlik^25^, Dorota Pasko^140^, Susan Walker^140^, Nick Parkinson^25^, Max Head Fourman^25^, Clark D Russell^25,141^, James Furniss^25^, Anne Richmond^24^, Elvina Gountouna^142^, David Harrison^138^, Bo Wang^24^, Yang Wu^143^, Alison Meynert^24^, Athanasios Kousathanas^140^, Loukas Moutsianas^140^, Zhijian Yang^144^, Ranran Zhai^144^, Chenqing Zheng^144^, Graeme Grimes^24^, Jonathan Millar^25^, Barbara Shih^25^, Marie Zechner^25^, Jian Yang^145,146^, Xia Shen^144,147,148^, Chris P. Ponting^2^, Albert Tenesa^2,25,147^, Kathy Rowan^138^, Andrew Law^25^, Veronique Vitart^24^, James F. Wilson^24,147^, J. Kenneth Baillie^24,25,26^.

**- Barts Health NHS Trust, London, UK**

D Collier^150^, S Wood^149^, A Zak^149^, C Borra^149^, M Matharu^149^, P May^149^, Z Alldis^149^, O Mitchelmore^149^, R Bowles^149^, A Easthorpe^149^, F Bibi^149^, I Lancoma-Malcolm^149^, J Gurasashvili^149^, J Pheby^149^, J Shiel^149^, M Bolton^149^, M Patel^149^, M Taylor^149^, O Zongo^149^, P Ebano^149^, P Harding^149^, R Astin-Chamberlain^149^, Y Choudhury^149^, A Cox^149^, D Kallon^149^, M Burton^149^, R Hall^149^, S Blowes^149^, Z Prime^149^, J Biddle^149^, O Prysyazhna^149^, T Newman^149^, C Tierney^149^, J Kassam^149^.

**- Guys and St Thomas’ Hospital, London, UK**

M Shankar-Hari^150^, M Ostermann^150^, S Campos^150^, A Bociek^150^, R Lim^150^, N Grau^150^, T O Jones^150^, C Whitton^150^, M Marotti^40^, G Arbane^150^.

**- James Cook University Hospital, Middlesburgh, UK**

S. Bonner^151^, K Hugill^151^, J Reid^151^.

**- The Royal Liverpool University Hospital, Liverpool, UK**

I Welters^152^, V Waugh^152^, K Williams^152^, D Shaw^152^, J Fernandez Roman^152^, M Lopez Martinez^152^, E Johnson^152^, A Waite^152^, B Johnson^152^, O Hamilton^152^, S Mulla^152^.

- **King’s College Hospital, London, UK**

M McPhail^153^, J Smith^153^.

- **Royal Infirmary of Edinburgh, Edinburgh, UK**

J K Baillie^23,24,25^, L Barclay^154^, D Hope^154^, C McCulloch^154^, L McQuillan^154^, S Clark^154^, J Singleton^154^, K Priestley^154^, N Rea^154^, M Callaghan^154^, R Campbell^154^, G Andrew^154^, L Marshall^154^.

- **John Radcliffe Hospital, Oxford, UK**

S McKechnie^155^, P Hutton^155^, A Bashyal ^155^, N Davidson^155^.

- **Addenbrooke’s Hospital, Cambridge, UK**

C Summers^155^, P Polgarova^155^, K Stroud^155^, N Pathan^155^, K Elston^155^, S Agrawal^155^.

- **Morriston Hospital, Swansea, UK**

C Battle^157^, L Newey^157^, T Rees^157^, R Harford^157^, E Brinkworth^157^, M Williams^157^, C Murphy^157^.

- **Ashford and St Peter’s Hospital, Surrey, UK**

I White^158^, M Croft^158^.

- **Royal Stoke University Hospital, Staffordshire, UK**

N Bandla^159^, M Gellamucho^159^, J Tomlinson^159^, H Turner^159^, M Davies^159^, A Quinn^159^, I Hussain^159^, C Thompson^159^, H Parker^159^, R Bradley^159^, R Griffiths^159^.

- **Queen Elizabeth Hospital, Birmingham, UK**

J. Scriven^160^, J Gill^160^.

- **Glasgow Royal Infirmary, Glasgow, UK**

A Puxty^161^, S Cathcart^161^, D Salutous^161^, L Turner^161^, K Duffy^161^, K Puxty^161^.

- **Kingston Hospital, Surrey, UK**

A Joseph^162^, R Herdman-Grant^162^, R Simms^162^, A Swain^162^, A Naranjo^162^, R Crowe^162^, K Sollesta^162^, A Loveridge^162^, D Baptista^162^, E Morino^162^.

- **The Tunbridge Wells Hospital and Maidstone Hospital, Kent, UK**

M Davey^163^, D Golden^163^, J Jones^163^.

- **North Middlesex University Hospital NHS trust, London, UK**

J Moreno Cuesta^164^, A Haldeos^164^, D Bakthavatsalam^164^, R Vincent^164^, M Elhassan^164^, K Xavier^164^, A Ganesan^164^, D Purohit M Abdelrazik^164^.

- **Bradford Royal Infirmary, Bradford, UK**

J Morgan^165^, L Akeroyd^165^, S Bano^165^, D Warren^165^, M Bromley^165^, K Sellick^165^, L Gurr^165^, B Wilkinson^165^, V Nagarajan^165^, P Szedlak^165^.

**- Blackpool Victoria Hospital, Blackpool, UK**

J Cupitt^166^, E Stoddard^166^, L Benham^166^, S Preston^166^, N Slawson^166^, Z Bradshaw^166^, J brown^166^, M Caswell^166^, SMelling^166^.

- **Countess of Chester Hospital, Chester, UK**

P Bamford^167^, M Faulkner^167^, K Cawley^167^, H Jeffrey^167^, E London^167^, H Sainsbury^167^, I Nagra^167^, F Nasir^167^, Ce Dunmore^167^, R Jones^167^, A Abraheem ^167^, M Al-Moasseb^167^, R Girach^167^.

- **Wythenshawe Hospital, Manchester, UK**

C Brantwood^168^, P Alexander ^168^, J Bradley-Potts^168^, S Allen^168^, T Felton^168^.

- **St George’s Hospital, London, UK**

S Manna^169^, S Farnell-Ward^169^, S Leaver^169^, J Queiroz^169^, E Maccacari^169^, D Dawson^169^, C Castro Delgado^169^, R Pepermans Saluzzio^169^, O Ezeobu^169^, L Ding^169^, C Sicat^169^, R Kanu^169^, G Durrant^169^, J Texeira^169^, A Harrison^169^, T Samakomva^169^.

**- Good Hope Hospital, Birmingham, UK**

J Scriven^170^, H Willis^170^, B Hopkins^170^, L Thrasyvoulou^170^.

- **Stepping Hill Hospital, Stockport, UK**

M Jackson^171^, A Zaki ^171^, C Tibke^171^, S Bennett^171^, W Woodyatt^171^, A Kent ^171^, E Goodwin^171^.

- **Manchester Royal Infirmary, Manchester, UK**

C Brandwood^172^, R Clark^172^, L Smith^172^.

- **Royal Alexandra Hospital, Paisley, UK**

K Rooney^173^, N Thomson^173^, N Rodden^173^, E Hughes^173^, D McGlynn^173^, C Clark^173^, P Clark^173^, L Abel^173^, R Sundaram^173^, L Gemmell^173^, M Brett^173^, J Hornsby^173^, P MacGoey^173^, R Price^173^, B Digby^173^, P O’Neil^173^, P McConnell^173^, P Henderson^173^.

- **Queen Elizabeth University Hospital, Glasgow, UK**

S Henderson^174^, M Sim^174^, S Kennedy-Hay^174^, C McParland^174^, L Rooney^174^, N Baxter^174^.

- **Queen Alexandra Hospital, Portsmouth, UK**

D Pogson^175^, S Rose^175^, Z Daly^175^, L Brimfield^175^.

- **BHRUT (Barking Havering) - Queens Hospital and King George Hospital, Essex, UK**

M K Phull^176^, M Hussain^176^, T Pogreban^176^, L Rosaroso^176^, E Salciute L Grauslyte^176^.

- **University College Hospital, London, UK**

D Brealey^177^, E Wraith^177^, N MacCallum^177^, G Bercades^177^, I Hass^177^, D Smyth^177^, A Reyes ^177^, G Martir^177^.

- **Royal Victoria Infirmary, Newcastle Upon Tyne, UK**

I D Clement^178^, K Webster^178^, C Hays^178^, A Gulati ^178^.

- **Western Sussex Hospitals, West Sussex, UK**

L Hodgson**^179^**, M Margarson^179^, R Gomez^179^, Y Baird^179^, Y Thirlwall^179^, L Folkes^179^, A Butler^179^, E Meadows^179^, S Moore^179^, D Raynard^179^, H Fox^179^, L Riddles^179^, K King^179^, S Kimber^179^, G Hobden^179^, A McCarthy^179^, V Cannons^179^, I Balagosa^179^, I Chadbourn^179^, A Gardner ^179^.

- **Salford Royal Hospital, Manchester, UK**

D Horner^180^, D McLaughlanv^180^, B Charles^180^, N Proudfoot^180^, T Marsden^180^, L Mc Morrow^180^, B Blackledge^180^, J Pendlebury^180^, A Harvey^180^, E Apetri^180^, C Basikolo^180^, L Catlow^180^, R Doonan^180^, K Knowles^180^, S Lee^180^, D Lomas^180^, C Lyons^180^, J Perez^180^, M Poulaka^180^, M Slaughter^180^, K Slevin^180^, M Taylor^180^, V Thomas^180^, D Walker^180^, J Harris^180^.

- **The Royal Oldham Hospital, Manchester, UK**

A Drummond^181^, R Tully^181^, J Dearden^181^, J Philbin^181^, S Munt^181^, C Rishton^181^, G O’Connor^181^, M Mulcahy^181^, E Dobson^181^, J Cuttler^181^, M Edward^181^.

- **Pinderfields General Hospital, Wakefield, UK**

A Rose^182^, B Sloan^182^, S Buckley^182^, H Brooke^182^, E Smithson^182^, R Charlesworth^182^, R Sandu^182^, M Thirumaran^182^, V Wagstaff^182^, J Cebrian Suarez^182^.

- **Basildon Hospital, Basildon, UK**

A Kaliappan^183^, M Vertue^183^, A Nicholson ^183^, J Riches^183^, A Solesbury^183^, L Kittridge^183^, M Forsey^183^, G Maloney^183^.

- **University Hospital of Wales, Cardiff, UK**

J Cole^184^, M Davies^184^, R Davies^184^, H Hill^184^, E Thomas^184^, A Williams ^184^, D Duffin^184^, B Player^184^.

**- Broomfield Hospital, Chelmsford, UK**

J Radhakrishnan^185^, S Gibson^185^, A Lyle ^185^, F McNeela^185^.

- **Royal Brompton Hospital, London, UK**

B Patel^186^, M Gummadi^186^, G Sloane^186^, N Dormand^186^, S Salmi^186^, Z Farzad^186^, D Cristiano^186^, K Liyanage^186^, V Thwaites^186^, M Varghese^186^.

- **Nottingham University Hospital, Nottingham, UK**

M Meredith^187^.

- **Royal Hallamshire Hospital and Northern General Hospital, Sheffield, UK**

G Mills^188^, J Willson^188^, K Harrington^188^, B Lenagh^188^, K Cawthron^188^, S Masuko^188^, A Raithatha^188^, K Bauchmuller^188^, N Ahmad^188^, J Barker^188^, Y Jackson^188^, F Kibutu^188^, S Bird^188^.

- **Royal Hampshire County Hospital, Hampshire, UK**

G Watson^189^, J Martin^189^, E Bevan^189^, C Wrey Brown^189^, D Trodd^189^.

- **Queens Hospital Burton, Burton-On-Trent, UK**

K English^190^, G Bell^190^, L Wilcox^190^, A Katary ^190^.

- **New Cross Hospital, Wolverhampton, UK**

S Gopal^191^, V Lake^191^, N Harris^191^, S Metherell^191^, E Radford^191^.

- **Heartlands Hospital, Birmingham, UK**

J Scriven^192^, F Moore^192^, H Bancroft^192^, J Daglish^192^, M Sangombe^192^, M Carmody^192^, J Rhodes^192^, M Bellamy^192^.

- **Walsall Manor Hospital, Walsall, UK**

A Garg^193^, A Kuravi ^193^, E Virgilio^193^, P Ranga^193^, J Butler^193^, L Botfield^193^, C Dexter^193^, J Fletcher^193^.

- **Stoke Mandeville Hospital, Buckinghamshire, UK**

P Shanmugasundaram^194^, G Hambrook^194^, I Burn^194^, K Manso^194^, D Thornton^194^, J Tebbutt^194^, R Penn^194^.

- **Sandwell General Hospital, Birmingham, UK**

J Hulme^195^, S Hussain^195^, Z Maqsood^195^, S Joseph^195^, J Colley^195^, A Hayes^195^, C Ahmed^195^, R Haque^195^, S Clamp^195^, R Kumar^195^, M Purewal^195^, B Baines^195^.

- **Royal Berkshire NHS Foundation Trust, Berkshire, UK**

M Frise^196^, N Jacques^196^, H Coles^196^, J Caterson^196^, S Gurung Rai^196^, M Brunton^196^, E Tilney^196^, L Keating^196^, A Walden^196^.

- **Charing Cross Hospital, St Mary’s Hospital and Hammersmith Hospital, London, UK**

D Antcliffe^197^, A Gordon ^197^, M Templeton^197^, R Rojo^197^, D Banach^197^, S Sousa Arias^197^, Z Fernandez^197^, P Coghlan^197^.

- **Dumfries and Galloway Royal Infirmary, Dumfries, UK**

D Williams^198^, C Jardine^198^.

- **Bristol Royal Infirmary, Bristol, UK**

J Bewley^199^, K Sweet^199^, L Grimmer^199^, R Johnson^199^, Z Garland^199^, B Gumbrill^199^.

- **Royal Sussex County Hospital, Brighton, UK**

C Phillips^200^, L Ortiz-Ruiz de Gordoa^200^, E Peasgood^200^.

- **Whiston Hospital, Prescot, UK**

A Tridente^201^, K Shuker S Greer^201^.

- **Royal Glamorgan Hospital, Cardiff, UK**

C Lynch^202^, C Pothecary^202^, L Roche^202^, B Deacon^202^, K Turner^202^, J Singh^202^, G Sera Howe^202^.

- **King’s Mill Hospital, Nottingham, UK**

P Paul^203^, M Gill^203^, I Wynter^203^, V Ratnam^203^, S Shelton^203^.

- **Fairfield General Hospital, Bury, UK**

J Naisbitt^204^, J Melville^204^.

- **Western General Hospital, Edinburgh, UK**

R Baruah^205^, S Morrison^205^.

- **Northwick Park Hospital, London, UK**

A McGregor^206^, V Parris^206^, M Mpelembue^206^, S Srikaran^206^, C Dennis^206^, A Sukha ^206^.

- **Royal Preston Hospital, Preston, UK**

A Williams^207^, M Verlande^207^.

- **Royal Derby Hospital, Derby, UK**

K Holding^208^, K Riches^208^, C Downes^208^, C Swan^208^.

- **Sunderland Royal Hospital, Sunderland, UK**

A Rostron^209^, A Roy ^209^, L Woods^209^, S Cornell^209^, F Wakinshaw^209^.

- **Royal Surrey County Hospital, Guildford, UK**

B Creagh-Brown^210^, H Blackman^210^, A Salberg^210^, E Smith^210^, S Donlon^210^, S Mtuwa^210^, N Michalak-Glinska^210^, S Stone^210^, C Beazley^210^, V Pristopan^210^.

- **Derriford Hospital, Plymouth, UK**

N Nikitas^211^, L Lankester^211^, C Wells^211^.

- **Croydon University Hospital, Croydon, UK**

A S Raj^212^, K Fletcher^212^, R Khade^212^, G Tsinaslanidis^212^.

- **Victoria Hospital, Kirkcaldy, UK**

M McMahon^213^, S Fowler^213^, A McGregor ^213^, T Coventry^213^.

- **Milton Keynes University Hospital, Milton Keynes, UK**

R Stewart^214^, L Wren^214^, E Mwaura^214^, L Mew^214^, A Rose^214^, D Scaletta^214^, F Williams^214^.

- **Barnsley Hospital, Barnsley, UK**

K Inweregbu^215^, A Nicholson^215^, N Lancaster^215^, M Cunningham^215^, A Daniels^215^, L Harrison^215^, S Hope^215^, S Jones^215^, A Crew^215^, G Wray^215^, J Matthews^215^, R Crawley^215^.

- **York Hospital, York, UK**

J Carter^216^, I Birkinshaw^216^, J Ingham^216^, Z Scott^216^, K Howard^216^, R Joy^216^, S Roche^216^.

- **University Hospital of North Tees, Stockton on Tees, UK**

M Clark^217^, S Purvis^217^.

- **University Hospital Wishaw, Wishaw, UK**

A Morrison^218^, D Strachan^218^, M Taylor^218^, S Clements^218^, K Black^218^.

**- Whittington Hospital, London, UK**

C Parmar^219^, A Altabaibeh ^219^, K Simpson^219^, L Mostoles^219^, K Gilbert^219^, L Ma^219^, A Alvaro ^219^.

**- Southmead Hospital, Bristol, UK**

M Thomas^220^, B Faulkner^220^, R Worner^220^, K Hayes^220^, E Gendall^220^, H Blakemore^220^, B Borislavova^220^, E Goff^220^.

**- The Royal Papworth Hospital, Cambridge, UK**

A Vuylsteke^221^, L Mwaura^221^, J Zamikula^221^, L Garner^221^, A Mitchell^221^, S Mepham^221^, L Cagova^221^, A Fofano^221^, H Holcombe^221^, K Praman^221^.

**- Royal Gwent Hospital, Newport, UK**

T Szakmany^222^, A E Heron ^222^, S Cherian^222^, S Cutler^222^, A Roynon-Reed^222^.

**- Norfolk and Norwich University hospital (NNUH), Norwich, UK**

G Randell^223^, K Convery^223^, K Stammers D Fottrell-Gould^223^, L Hudig^223^, J Keshet-price^223^.

**- Great Ormond St Hospital and UCL Great Ormond St Institute of Child Health NIHR Biomedical Research Centre, London, UK**

M Peters^224^, L O’Neill^224^, S Ray^224^, H Belfield^224^, T McHugh^224^, G Jones^224^, O Akinkugbe^224^, A Tomas^224^, E Abaleke^224^, E Beech^224^, H Meghari^224^, S Yussuf^224^, A Bamford ^224^.

**- Airedale General Hospital, Keighley, UK**

B Hairsine^225^, E Dooks^225^, F Farquhar^225^, S Packham^225^, H Bates^225^, C McParland^225^, L Armstrong ^225^.

**- Aberdeen Royal Infirmary, Aberdeen, UK**

C Kaye^226^, A Allan ^226^, J Medhora^226^, J Liew^226^, A Botello^226^, F Anderson^226^.

**- Southampton General Hospital, Southampton, UK**

R Cusack^227^, H Golding^227^, K Prager^227^, T Williams^227^, S Leggett^227^, K Golder^227^, M Male^227^, O Jones^227^, K Criste^227^, M Marani^227^.

**- Russell’s Hall Hospital, Dudley, UK**

V. Anumakonda^228^, V Amin ^228^, K Karthik^228^, R Kausar^228^, E Anastasescu^228^, K Reid^228^, M. Jacqui^228^.

**- Rotherham General Hospital, Rotherham, UK**

A Hormis^229^, R Walker^229^, D Collier^229^.

**- North Manchester General Hospital, Manchester, UK**

T Duncan^230^, A Uriel ^230^, A Ustianowski ^230^, H T-Michael^230^, M Bruce^230^, K Connolly^230^, K Smith^230^.

**- Basingstoke and North Hampshire Hospital, Basingstoke, UK**

R Partridge^231^, D Griffin^231^, M McDonald^231^, N Muchenje^231^.

**- Royal Free Hospital, London, UK**

D Martin^232^, H Filipe^232^, C Eastgate^232^, C Jackson^232^.

**- Hull Royal Infirmary, Hull, UK**

A Gratrix^233^, L Foster^233^, V Martinson^233^, E Stones^233^, Caroline Abernathy^233^, P Parkinson^233^.

**- Harefield Hospital, London, UK**

A Reed^234^, C Prendergast^234^, P Rogers^234^, M Woodruff^234^, R Shokkar^234^, S Kaul^234^, A Barron ^234^, C Collins^234^.

**- Chesterfield Royal Hospital Foundation Trust, Chesterfield, UK**

S Beavis^235^, A Whileman ^235^, K Dale^235^, J Hawes^235^, K Pritchard^235^, R Gascoyne^235^, L Stevenson^235^.

**- Barnet Hospital, London, UK**

R Jha^236^, L Lim^236^, V Krishnamurthy^236^.

**- Aintree University Hospital, Liverpool, UK**

R Parker^237^, I Turner-Bone^237^, L Wilding^237^, A Reddy ^237^.

**- St James’s University Hospital and Leeds General Infirmary, Leeds, UK**

S Whiteley^238^, E Wilby^238^, C Howcroft^238^, A Aspinwall ^238^, S Charlton^238^, B Ogg^238^.

**- Glan Clwyd Hospital, Bodelwyddan, UK**

D Menzies^239^, R Pugh^239^, E Allan^239^, R Lean^239^, F Davies^239^, J Easton^239^, X Qiu^239^, S Kumar^239^, K Darlington^239^.

**- University Hospital Crosshouse, Kilmarnock, UK**

G Houston^240^, P O’Brien^240^, T Geary^240^, J Allan^240^, A Meikle^240^.

**- Royal Bolton Hospital, Bolton, UK**

G Hughes^241^, M Balasubramaniam^241^, S Latham^241^, E McKenna^241^, R Flanagan^241^.

**- Princess of Wales Hospital, Llantrisant, UK**

S Sathe^242^, E Davies^242^, L Roche^242^.

**- Pilgrim Hospital, Lincoln, UK**

M Chablani^243^, A Kirkby ^243^, K Netherton^243^, S Archer^243^.

**- Northumbria Healthcare NHS Foundation Trust, North Shields, UK**

B Yates^244^, C Ashbrook-Raby^244^.

**- Ninewells Hospital, Dundee, UK**

S Cole^245^, M Casey^245^, L Cabrelli^245^, S Chapman^245^, M Casey^245^, P Austin^245^, A Hutcheon^245^, C Whyte^245^, C Almaden-Boyle^245^.

**- Lister Hospital, Stevenage, UK**

N Pattison^246^, C Cruz^246^.

**- Bedford Hospital, Bedford, UK**

A Vochin^247^, H Kent^247^, A Thomas^247^, S Murdoch^247^, B David^247^, M Penacerrada^247^, G Lubimbi^247^, V Bastion^247^, R Wulandari^247^, J Valentine^247^, D Clarke^247^.

**- Royal United Hospital, Bath, UK**

A Serrano-Ruiz^248^, S Hierons^248^, L Ramos^248^, C Demetriou^248^, S Mitchard^248^, K White^248^.

**- Royal Bournemouth Hospital, Bournemouth, UK**

N White^249^, S Pitts^249^, D Branney^249^, J Frankham^249^.

**- The Great Western Hospital, Swindon, UK**

M Watters^250^, H Langton^250^, R Prout^250^.

**- Watford General Hospital, Watford, UK**

V Page^251^, T Varghes^251^.

**- University Hospital North Durham, Darlington, UK**

A Cowton^252^, A Kay ^252^, K Potts^252^, M Birt^252^, M Kent^252^, A Wilkinson^252^.

**- Tameside General Hospital, Ashton Under Lyne, UK**

E Jude^253^, V Turner^253^, H Savill^253^, J McCormick^253^, M Clark^253^, M Coulding^253^, S Siddiqui^253^, O Mercer^253^, H Rehman^253^, D Potla^253^.

**- Princess Royal Hospital, Telford and Royal Shrewsbury Hospital, Shrewsbury, UK**

N Capps^254^, D Donaldson^254^, J Jones^254^, H Button^254^, T Martin^254^, K Hard^254^, A Agasou^254^, L Tonks^254^, T Arden^254^, P Boyle^254^, M Carnahan^254^, J Strickley^254^, C Adams^254^, D Childs^254^, R Rikunenko^254^, M Leigh^254^, M Breekes^254^, R Wilcox^254^, A Bowes^254^, H Tiveran^254^, F Hurford^254^, J Summers^254^, A Carter^254^, Y Hussain^254^, L Ting^254^, A Javaid^254^, N Motherwell^254^, H Moore^254^, H Millward^254^, S Jose^254^, N Schunki^254^, A Noakes ^254^, C Clulow^254^

**- Arrowe Park Hospital, Wirral, UK**

G Sadera^255^, R Jacob^255^, C Jones^255^

**- The Queen Elizabeth Hospital, King’s Lynn, UK**

M Blunt^256^, Z Coton^256^, H Curgenven^256^, S Mohamed Ally^256^, K Beaumont^256^, M Elsaadany^256^, K Fernandes^256^, I Ali Mohamed Ali^256^, H Rangarajan^256^, V Sarathy^256^, S Selvanayagam^256^, D Vedage^256^, M White^256^

**- Royal Blackburn Teaching Hospital, Blackburn, UK**

M Smith^257^, N Truman^257^, S Chukkambotla^257^, S Keith^257^, J Cockerill-Taylor^257^, J Ryan-Smith^257^, R Bolton^257^, P Springle^257^, J Dykes^257^, J Thomas^257^, M Khan^257^, M T Hijazi^257^, E Massey^257^, G Croston^257^

**- Poole Hospital, Poole, UK**

H Reschreite r^258^, J Camsooksai^258^, S Patch^258^, S Jenkins^258^, C Humphrey^258^, B Wadams^258^, J Camsooksai^258^.

**- Medway Maritime Hospital, Gillingham, UK**

N Bhatia^259^, M Msiska^259^, O Adanini^259^.

**- Warwick Hospital, Warwick, UK**

B Attwood^260^, P Parsons^260^.

**- The Royal Marsden Hospital, London, UK**

K Tatham^261^, S Jhanji^261^, E Black^261^, A Dela Rosa ^261^, R Howle^261^, B Thomas^261^, T Bemand^261^, R Raobaikady^261^

**- The Princess Alexandra Hospital, Harlow, UK**

R Saha^262^, N Staines^262^, A Daniel ^262^, J Finn^262^.

**- Musgrove Park Hospital, Taunton, UK**

J Hutter^263^, P Doble^263^, C Shovelton^263^, C Pawley^263^.

**- George Eliot Hospital NHS Trust, Nuneaton, UK**

T Kannan^264^, M Hill^264^.

**- East Surrey Hospital, Redhill, UK**

E Combes^265^, S Monnery^265^, T Joefield^265^.

**- West Middlesex Hospital, Isleworth, UK**

M Popescu^266^, M Thankachen^266^, M Oblak^266^.

**- Warrington General Hospital, Warrington, UK**

J Little^267^, S McIvor^267^, A Brady ^267^, H Whittle^267^, H Prady^267^, R Chan^267^

**- Southport and Formby District General Hospital, Ormskirk, UK**

A Ahmed ^268^, A Morris ^268^.

**- Royal Devon and Exeter Hospital, Exeter, UK**

C Gibson^269^, E Gordon^269^, S Keenan^269^, H Quinn^269^, S Benyon^269^, S Marriott^269^, L Zitter^269^, L Park^269^, K Baines^269^

**- Macclesfield District General Hospital, Macclesfield, UK**

M Lyons^270^, M Holland^270^, N Keenan^270^, M Young^270^.

**- Borders General Hospital, Melrose, UK**

S Garrioch^271^, J Dawson^271^, M Tolson^271^.

**- Birmingham Children’s Hospital, Birmingham, UK**

B Scholefield^272^, R Bi^272^.

**- William Harvey Hospital, Ashford, UK**

N Richardson^273^, N Schumacher^273^, T Cosier^273^, G Millen^273^

**- Royal Lancaster Infirmary, Lancaster, UK**

A Higham^274^, K Simpson^274^

**- Queen Elizabeth the Queen Mother Hospital, Margate, UK**

S Turki^275^, L Allen^275^, N Crisp^275^, T Hazleton^275^, A Knight^275^, J Deery^275^, C Price^275^, S Turney^275^, S Tilbey^275^, E Beranova^275^

**- Liverpool Heart and Chest Hospital, Liverpool, UK**

D Wright^276^, L Georg^276^, S Twiss^276^.

**- Darlington Memorial Hospital, Darlington, UK**

A Cowton^277^, S Wadd^277^, K Postlethwaite^277^.

**- Southend University Hospital, Westcliff-on-Sea, UK**

P Gondo^278^, B Masunda^278^, A Kayani ^278^, B Hadebe^278^

**- Raigmore Hospital, Inverness, UK**

J Whiteside^279^, R Campbell^279^, N Clarke^279^

**- Salisbury District Hospital, Salisbury, UK**

P Donnison^280^, F Trim^280^, I Leadbitter^280^

**- Peterborough City Hospital, Peterborough, UK**

D Butcher^281^, S O’Sullivan^281^

**- Ipswich Hospital, Ipswich, UK**

B Purewal^282^, S Bell^282^, V Rivers’^282^

**- Hereford County Hospital, Hereford, UK**

R O’Leary^283^, J Birch^283^, E Collins^283^, S Anderson^283^, K Hammerton^283^, E Andrews^283^

**- Furness General Hospital, Barrow-in-Furness, UK**

A Higham^284^, K Burns^284^

**- Forth Valley Royal Hospital, Falkirk, UK**

I Edmond^285^, D Salutous^285^, A Todd ^285^, J Donnachie^285^, P Turner^285^, L Prentice^285^, L Symon^285^, N Runciman^285^, F Auld^285^

**- Torbay Hospital, Torquay, UK**

M Halkes^286^, P Mercer^286^, L Thornton^286^

**- St Mary’s Hospital, Newport, UK**

G Debreceni^287^, J Wilkins^287^, A Brown ^287^, V Crickmore^287^.

**- Royal Manchester Children’s Hospital, Manchester, UK**

G Subramanian^288^, R Marshall^288^, C Jennings^288^, M Latif^288^, L Bunni^288^

**- Royal Cornwall Hospital, Truro, UK**

M Spivey^289^, S Bean^289^, K Burt^289^

**- Queen Elizabeth Hospital Gateshead, Gateshead, UK**

V Linnett^290^, J Ritzema^290^, A Sanderson ^290^, W McCormick^290^, M Bokhari^290^

**- Kent & Canterbury Hospital, Canterbury, UK**

R Kapoor^291^, D Loader^291^

**- James Paget University Hospital NHS Trust, Great Yarmouth, UK**

A Ayers ^292^, W Harrison^292^, J North^292^

**- Darent Valley Hospital, Dartford, UK**

Z Belagodu^293^, R Parasomthy^293^, O Olufuwa^293^, A Gherman ^293^, B Fuller^293^, C Stuart^293^

**- The Alexandra Hospital, Redditch and Worcester Royal Hospital, Worcester, UK**

O Kelsall^294^, C Davis^294^, L Wild^294^, H Wood^294^, J Thrush^294^, A Durie^294^, K Austin’^294^, K Archer^294^, P Anderson^294^, C Vigurs^294^

**- Ysbyty Gwynedd, Bangor, UK**

C Thorpe^295^, A Thomas ^295^, E Knights^295^, N Boyle^295^, A Price^295^

**- Yeovil Hospital, Yeovil, UK**

A Kubisz-Pudelko^296^, D Wood^296^, A Lewis ^296^, S Board^296^, L Pippard^296^, J Perry^296^, K Beesley^296^

**- University Hospital Hairmyres, East Kilbride, UK**

A Rattray^297^, M Taylor^297^, E Lee^297^, L Lennon^297^, K Douglas^297^, D Bell^297^, R Boyle^297^, L Glass^297^

**- Scunthorpe General Hospital, Scunthorpe, UK**

M Nauman Akhtar^298^, K Dent^298^, D Potoczna^298^, S Pearson^298^, E Horsley^298^, S Spencer^298^

**- Princess Royal Hospital Brighton, West Sussex, UK**

C Phillips^299^, D Mullan^299^, D Skinner^299^, J Gaylard^299^, L Ortiz-Ruizdegordoa^299^.

**- Lincoln County Hospital, Lincoln, UK**

R Barber^300^, C Hewitt^300^, A Hilldrith ^300^, S Shepardson^300^, M Wills^300^, K Jackson-Lawrence^300^

**- Homerton University Hospital, London, UK**

A Gupta^301^, A Easthope ^301^, E Timlick^301^, C Gorman^301^.

**- Glangwili General Hospital, Camarthen, UK**

I Otaha^302^, A Gales ^302^, S Coetzee^302^, M Raj^302^, M Peiu^302^

**- Ealing Hospital, Southall, UK**

V Parris^303^, S Quaid^303^, E Watson^303^

**- Scarborough General Hospital, Scarborough, UK**

K Elliott^304^, J Mallinson^304^, B Chandler^304^, A Turnbull ^304^

**- Royal Albert Edward Infirmary, Wigan, UK**

A Quinn^305^, C Finch^305^, C Holl^305^, J Cooper^305^, A Evans^305^.

**- Queen Elizabeth Hospital, Woolwich, London, UK**

W Khaliq^306^, A Collins ^306^, E Treus Gude^306^

**- North Devon District Hospital, Barnstaple, UK**

N Love^307^, L van Koutrik^307^, J Hunt^307^, D Kaye^307^, E Fisher^307^, A Brayne ^307^, V Tuckey^307^, P Jackson^307^, J Parkin^307^

**- National Hospital for Neurology and Neurosurgery, London, UK**

D Brealey^308^, E Raith^308^, A Tariq ^308^, H Houlden^308^, A Tucci^308^, J Hardy^308^, E Moncur^308^.

**- Eastbourne District General Hospital, East Sussex, UK and Conquest Hospital, East Sussex, UK**

J Highgate^309^, A Cowley ^309^

**- Diana Princess of Wales Hospital, Grimsby, UK**

A Mitra^310^, R Stead^310^, T Behan^310^, C Burnett^310^, M Newton^310^, E Heeney^310^, R Pollard^310^, J Hatton^310^

**- The Christie NHS Foundation Trust, Manchester, UK**

A Patel^311^, V Kasipandian^311^, S Allibone^311^, R M Genetu^311^

**- Prince Philip Hospital, Lianelli, UK**

I Otahal^312^, L O’Brien^312^, Z Omar^312^, E Perkins^312^, K Davies^312^

**- Prince Charles Hospital, Merthyr Tydfil, UK**

D Tetla^313^, C Pothecary^313^, B Deacon^313^

**- Golden Jubilee National Hospital, Clydebank, UK**

B Shelley^314^, V Irvine^314^

**- Dorset County Hospital, Dorchester, UK**

S Williams^315^, P Williams^315^, J Birch^315^, J Goodsell^315^, R Tutton^315^, L Bough^315^, B Winter-Goodwin^315^

**- Calderdale Royal Hospital, Halifax, UK**

R Kitson^316^, J Pinnell^316^, A Wilson^316^, T Nortcliffe^316^, T Wood^316^, M Home^316^, K Holdroyd^316^, M Robinson^316^, R Shaw^316^, J Greig^316^, M Brady^316^, A Haigh^316^, L Matupe^316^, M Usher^316^, S Mellor^316^, S Dale^316^, L Gledhill^316^, L Shaw^316^, G Turner^316^, D Kelly^316^, B Anwar^316^, H Riley^316^, H Sturgeon^316^, A Ali ^316^, L Thomis^316^, D Melia^316^, A Dance ^316^, K Hanson^316^

**- West Suffolk Hospital, Suffolk, UK**

S Humphreys^317^, I Frost^317^, V Gopal^317^, J Godden^317^, A Holden^317^, S Swann^317^.

**- West Cumberland Hospital, Whitehaven, UK**

T Smith^318^, M Clapham^318^, U Poultney^318^, R Harper^318^, P Rice^318^.

**- University Hospital Lewisham, London, UK**

W Khaliq^319^, R Reece-Anthony^319^, B Gurung^319^.

**- St John’s Hospital Livingston, Livingston, UK**

S Moultrie^320^, M Odam^320^

**- Sheffield Children’s Hospital, Sheffield, UK**

A Mayer^321^, A Bellini ^321^, A Pickard ^321^, J Bryant^321^, N Roe^321^, J Sowter^321^

**- Hinchingbrooke Hospital, Huntingdon, UK**

D Butcher^322^, K Lang^322^, J Taylor^322^.

**- Glenfield Hospital, Leicester, UK**

P Barry^323^

**- Bronglais General Hospital, Aberystwyth, UK**

M Hobrok^324^, H Tench^324^, R Wolf-Roberts^324^, H McGuinness^324^, R Loosley^324^

**- Alder Hey Children’s Hospital, Liverpool, UK**

D Hawcutt^325^, L Rad^325^, L O’Malley^325^, P Saunderson^325^, G Seddon^325^, T Anderson ^325^, N Rogers^325^

**- University Hospital Monklands, Airdrie, UK**

J Ruddy^326^, M Harkins^326^, M Taylor^326^, C Beith^326^, A McAlpine^326^, L Ferguson^326^, P Grant^326^, S MacFadyen^326^, M McLaughlin^326^, T Baird^326^, S Rundell^326^, L Glass^326^, B Welsh^326^, R Hamill^326^, F Fisher^326^.

**- Cumberland Infirmary, Carlisle, UK**

T Smith^327^, J Gregory^327^, A Brown ^327^

122) Centre for Tropical Medicine and Global Health, Nuffield Department of Medicine, University of Oxford, Old Road Campus, Roosevelt Drive, Oxford, OX3 7FZ, UK

123) Wellcome Centre for Human Genetics, University of Oxford, Oxford, UK

124) UCL Centre for Human Health and Performance, London, W1T 7HA, UK

125) Department of Critical Care Medicine, Queen’s University and Kingston Health Sciences Centre, Kingston, ON, Canada

126) Department of Anaesthesia and Intensive Care, The Chinese University of Hong Kong, Prince of Wales Hospital, Hong Kong, China

127) Clinical Research Centre at St Vincent’s University Hospital, University College Dublin, Dublin, Ireland

128) Department of Medicine, University of Cambridge, Cambridge, UK

129) William Harvey Research Institute, Barts and the London School of Medicine and Dentistry, Queen Mary University of London, London EC1M 6BQ, UK

130) NIHR Health Protection Research Unit for Emerging and Zoonotic Infections, Institute of Infection, Veterinary and Ecological Sciences University of Liverpool, Liverpool, L69 7BE, UK

131) Respiratory Medicine, Alder Hey Children’s Hospital, Institute in The Park, University of Liverpool, Alder Hey Children’s Hospital, Liverpool, UK

132) National Heart and Lung Institute, Imperial College London, London, UK

133) Imperial College Healthcare NHS Trust: London, London, UK

134) Department of Intensive Care Medicine, Guy’s and St. Thomas NHS Foundation Trust, London, UK

135) MRC-University of Glasgow Centre for Virus Research, Institute of Infection, Immunity and Inflammation, College of Medical, Veterinary and Life Sciences, University of Glasgow, Glasgow, UK

136) Wellcome-Wolfson Institute for Experimental Medicine, Queen’s University Belfast, Belfast, Northern Ireland, UK

137) Department of Intensive Care Medicine, Royal Victoria Hospital, Belfast, Northern Ireland, UK

138) Intensive Care National Audit & Research Centre, London, UK

139) Edinburgh Clinical Research Facility, Western General Hospital, University of Edinburgh, EH4 2XU, UK

140) Genomics England, London, UK

141) Centre for Inflammation Research, The Queen’s Medical Research Institute, University of Edinburgh, 47 Little France Crescent, Edinburgh, UK

142) Centre for Genomic and Experimental Medicine, Institute of Genetics and Molecular Medicine, University of Edinburgh, Western General Hospital, Crewe Road, Edinburgh, EH4 2XU, UK

143) Institute for Molecular Bioscience, The University of Queensland, Brisbane, Australia

144) Biostatistics Group, School of Life Sciences, Sun Yat-sen University, Guangzhou, China

145) School of Life Sciences, Westlake University, Hangzhou, Zhejiang 310024, China

146) Westlake Laboratory of Life Sciences and Biomedicine, Hangzhou, Zhejiang 310024, China

147) Centre for Global Health Research, Usher Institute of Population Health Sciences and Informatics, Teviot Place, Edinburgh EH8 9AG, UK

148) Department of Medical Epidemiology and Biostatistics, Karolinska Institutet, Stockholm, Sweden

149) Barts Health NHS Trust, London, UK

150) Guys and St Thomas’ Hospital, London, UK

151) James Cook University Hospital, Middlesburgh, UK

152) The Royal Liverpool University Hospital, Liverpool, UK

153) King’s College Hospital, London, UK

154) Royal Infirmary of Edinburgh, Edinburgh, UK

155) John Radcliffe Hospital, Oxford, UK

156) Addenbrooke’s Hospital, Cambridge, UK

157) Morriston Hospital, Swansea, UK

158) Ashford and St Peter’s Hospital, Surrey, UK

159) Royal Stoke University Hospital, Staffordshire, UK

160) Queen Elizabeth Hospital, Birmingham, UK

161) Glasgow Royal Infirmary, Glasgow, UK

162) Kingston Hospital, Surrey, UK

163) The Tunbridge Wells Hospital and Maidstone Hospital, Kent, UK

164) North Middlesex University Hospital NHS trust, London, UK

165) Bradford Royal Infirmary, Bradford, UK

166) Blackpool Victoria Hospital, Blackpool, UK

167) Countess of Chester Hospital, Chester, UK

168) Wythenshawe Hospital, Manchester, UK

169) St George’s Hospital, London, UK

170) Good Hope Hospital, Birmingham, UK

171) Stepping Hill Hospital, Stockport, UK

172) Manchester Royal Infirmary, Manchester, UK

173) Royal Alexandra Hospital, Paisley, UK

174) Queen Elizabeth University Hospital, Glasgow, UK

175) Queen Alexandra Hospital, Portsmouth, UK

176) BHRUT (Barking Havering) - Queens Hospital and King George Hospital, Essex, UK

177) University College Hospital, London, UK

178) Royal Victoria Infirmary, Newcastle Upon Tyne, UK

179) Western Sussex Hospitals, West Sussex, UK

180) Salford Royal Hospital, Manchester, UK

181) The Royal Oldham Hospital, Manchester, UK

182) Pinderfields General Hospital, Wakefield, UK

183) Basildon Hospital, Basildon, UK

184) University Hospital of Wales, Cardiff, UK

185) Broomfield Hospital, Chelmsford, UK

186) Royal Brompton Hospital, London, UK

187) Nottingham University Hospital, Nottingham, UK

188) Royal Hallamshire Hospital and Northern General Hospital, Sheffield, UK

189) Royal Hampshire County Hospital, Hampshire, UK

190) Queens Hospital Burton, Burton-On-Trent, UK

191) New Cross Hospital, Wolverhampton, UK

192) Heartlands Hospital, Birmingham, UK

193) Walsall Manor Hospital, Walsall, UK

194) Stoke Mandeville Hospital, Buckinghamshire, UK

195) Sandwell General Hospital, Birmingham, UK

196) Royal Berkshire NHS Foundation Trust, Berkshire, UK

197) Charing Cross Hospital, St Mary’s Hospital and Hammersmith Hospital, London, UK

198) Dumfries and Galloway Royal Infirmary, Dumfries, UK

199) Bristol Royal Infirmary, Bristol, UK

200) Royal Sussex County Hospital, Brighton, UK

201) Whiston Hospital, Prescot, UK

202) Royal Glamorgan Hospital, Cardiff, UK

203) King’s Mill Hospital, Nottingham, UK

204) Fairfield General Hospital, Bury, UK

205) Western General Hospital, Edinburgh, UK

206) Northwick Park Hospital, London, UK

207) Royal Preston Hospital, Preston, UK

208) Royal Derby Hospital, Derby, UK

209) Sunderland Royal Hospital, Sunderland, UK

210) Royal Surrey County Hospital, Guildford, UK

211) Derriford Hospital, Plymouth, UK

212) Croydon University Hospital, Croydon, UK

213) Victoria Hospital, Kirkcaldy, UK

214) Milton Keynes University Hospital, Milton Keynes, UK

215) Barnsley Hospital, Barnsley, UK

216) York Hospital, York, UK

217) University Hospital of North Tees, Stockton on Tees, UK

218) University Hospital Wishaw, Wishaw, UK

219) Whittington Hospital, London, UK

220) Southmead Hospital, Bristol, UK

221) The Royal Papworth Hospital, Cambridge, UK

222) Royal Gwent Hospital, Newport, UK

223) Norfolk and Norwich University hospital (NNUH), Norwich, UK

224) Great Ormond St Hospital and UCL Great Ormond St Institute of Child Health NIHR Biomedical Research Centre, London, UK

225) Airedale General Hospital, Keighley, UK

226) Aberdeen Royal Infirmary, Aberdeen, UK

227) Southampton General Hospital, Southampton, UK

228) Russell’s Hall Hospital, Dudley, UK

229) Rotherham General Hospital, Rotherham, UK

230) North Manchester General Hospital, Manchester, UK

231) Basingstoke and North Hampshire Hospital, Basingstoke, UK

232) Royal Free Hospital, London, UK

233) Hull Royal Infirmary, Hull, UK

234) Harefield Hospital, London, UK

235) Chesterfield Royal Hospital Foundation Trust, Chesterfield, UK

236) Barnet Hospital, London, UK

237) Aintree University Hospital, Liverpool, UK

238) St James’s University Hospital and Leeds General Infirmary, Leeds, UK

239) Glan Clwyd Hospital, Bodelwyddan, UK

240) University Hospital Crosshouse, Kilmarnock, UK

241) Royal Bolton Hospital, Bolton, UK

242) Princess of Wales Hospital, Llantrisant, UK

243) Pilgrim Hospital, Lincoln, UK

244) Northumbria Healthcare NHS Foundation Trust, North Shields, UK

245) Ninewells Hospital, Dundee, UK

246) Lister Hospital, Stevenage, UK

247) Bedford Hospital, Bedford, UK

248) Royal United Hospital, Bath, UK

249) Royal Bournemouth Hospital, Bournemouth, UK

250) The Great Western Hospital, Swindon, UK

251) Watford General Hospital, Watford, UK

252) University Hospital North Durham, Darlington, UK

253) Tameside General Hospital, Ashton Under Lyne, UK

254) Princess Royal Hospital Shrewsbury and Royal Shrewsbury Hospital, Shrewsbury, UK

255) Arrowe Park Hospital, Wirral, UK

256) The Queen Elizabeth Hospital, King’s Lynn, UK

257) Royal Blackburn Teaching Hospital, Blackburn, UK

258) Poole Hospital, Poole, UK

259) Medway Maritime Hospital, Gillingham, UK

260) Warwick Hospital, Warwick, UK

261) The Royal Marsden Hospital, London, UK

262) The Princess Alexandra Hospital, Harlow, UK

263) Musgrove Park Hospital, Taunton, UK

264) George Eliot Hospital NHS Trust, Nuneaton, UK

265) East Surrey Hospital, Redhill, UK

266) West Middlesex Hospital, Isleworth, UK

267) Warrington General Hospital, Warrington, UK

268) Southport and Formby District General Hospital, Ormskirk, UK

269) Royal Devon and Exeter Hospital, Exeter, UK

270) Macclesfield District General Hospital, Macclesfield, UK

271) Borders General Hospital, Melrose, UK

272) Birmingham Children’s Hospital, Birmingham, UK

273) William Harvey Hospital, Ashford, UK

274) Royal Lancaster Infirmary, Lancaster, UK

275) Queen Elizabeth the Queen Mother Hospital, Margate, UK

276) Liverpool Heart and Chest Hospital, Liverpool, UK

277) Darlington Memorial Hospital, Darlington, UK

278) Southend University Hospital, Westcliff-on-Sea, UK

279) Raigmore Hospital, Inverness, UK

280) Salisbury District Hospital, Salisbury, UK

281) Peterborough City Hospital, Peterborough, UK

282) Ipswich Hospital, Ipswich, UK

283) Hereford County Hospital, Worcester, UK

284) Furness General Hospital, Barrow-in-Furness, UK

285) Forth Valley Royal Hospital, Falkirk, UK

286) Torbay Hospital, Torquay, UK

287) St Mary’s Hospital, Newport, UK

288) Royal Manchester Children’s Hospital, Manchester, UK

289) Royal Cornwall Hospital, Truro, UK

290) Queen Elizabeth Hospital Gateshead, Gateshead, UK

291) Kent & Canterbury Hospital, Canterbury, UK

292) James Paget University Hospital NHS Trust, Great Yarmouth, UK

293) Darent Valley Hospital, Dartford, UK

294) The Alexandra Hospital, Redditch and Worcester Royal Hospital, Worcester, UK

295) Ysbyty Gwynedd, Bangor, UK

296) Yeovil Hospital, Yeovil, UK

297) University Hospital Hairmyres, East Kilbride, UK

298) Scunthorpe General Hospital, Scunthorpe, UK

299) Princess Royal Hospital Brighton, West Sussex, UK

300) Lincoln County Hospital, Lincoln, UK

301) Homerton University Hospital, London, UK

302) Glangwili General Hospital, Camarthen, UK

303) Ealing Hospital, Southall, UK

304) Scarborough General Hospital, Scarborough, UK

305) Royal Albert Edward Infirmary, Wigan, UK

306) Queen Elizabeth Hospital, Woolwich, London, UK

307) North Devon District Hospital, Barnstaple, UK

308) National Hospital for Neurology and Neurosurgery, London, UK

309) Eastbourne District General Hospital, East Sussex, UK and Conquest Hospital, East Sussex, UK

310) Diana Princess of Wales Hospital, Grimsby, UK

311) The Christie NHS Foundation Trust, Manchester, UK

312) Prince Philip Hospital, Lianelli, UK

313) Prince Charles Hospital, Merthyr Tydfil, UK

314) Golden Jubilee National Hospital, Clydebank, UK

315) Dorset County Hospital, Dorchester, UK

316) Calderdale Royal Hospital, Halifax, UK

317) West Suffolk Hospital, Suffolk, UK

318) West Cumberland Hospital, Whitehaven, UK

319) University Hospital Lewisham, London, UK

320) St John’s Hospital Livingston, Livingston, UK

321) Sheffield Children’s Hospital, Sheffield, UK

322) Hinchingbrooke Hospital, Huntingdon, UK

323) Glenfield Hospital, Leicester, UK

324) Bronglais General Hospital, Aberystwyth, UK

325) Alder Hey Children’s Hospital, Liverpool, UK

326) University Hospital Monklands, Airdrie, UK

327) Cumberland Infirmary, Carlisle, UK

**-German COVID-19 OMICS Initiative (DeCOI) - Host genetics subgroup**

Axel Schmidt^20^, Kerstin U. Ludwig^20^, Selina Rolker^20^, Markus M. Nöthen^20^, Julia Fazaal^20^, Verena Keitel^329^, Björn Jensen^328^, Torsten Feldt^328^, Lisa Knopp^328^, Julia Schröder^21^, Carlo Maj^329^, Fabian Brand^329^, Marc M. Berger^329^, Thorsten Brenner^330^, Anke Hinney^331^, Oliver Witzke^332^, Robert Bals^333^, Christian Herr^333^, Nicole Ludwig^334^, Jörn Walter^335^, Jochen Schneider^336^, Johanna Erber^336^, Christoph D. Spinner^336,337^, Clemens M. Wendtner^338^, Christof Winter^339^, Ulrike Protzer^340^, Nicolas Casadei^341^, Stephan Ossowski^342^, Olaf H. Riess^343^, Eva C. Schulte^21,22.23^

328) Department of Gastroenterology, Hepatology and Infectious Diseases, University Hospital Duesseldorf, Medical Faculty Heinrich Heine University, Duesseldorf, Germany

329) Institute of Genomic Statistics and Bioinformatics, University Hospital Bonn, Medical Faculty University of Bonn, Venusberg-Campus 1, Bonn Germany

330) Department of Anesthesiology and Intensive Care Medicine, University Hospital Essen, University Duisburg-Essen, Essen, Germany

331) Department of Child and Adolescent Psychiatry, University Hospital Essen, University of Duisburg-Essen, Essen, Germany

332) Department of Infectious Diseases, University Hospital Essen, University Duisburg-Essen, Essen, Germany

333) Department of Internal Medicine V - Pneumology, Allergology and Intensive Care Medicine, University Hospital Saarland, Homburg/Saar, Germany

334) Center of Human and Molecular Biology, Department of Human Genetics, University Hospital Saarland, Homburg/Saar, Germany

335) Department of Genetics & Epigenetics, Saarland University, Saarbrücken, Germany

336) Department of Internal Medicine II, School of Medicine, University Hospital Rechts der Isar, Technical University of Munich, Munich, Germany.

337) German Center for Infection Research (DZIF), Partner Site Munich, Munich, Germany.

338) Department of Hematology, Oncology, Immunology, Palliative Medicine, Infectious Diseases and Tropical Medicine, Munich Clinic Schwabing, Academic Teaching Hospital (LMU), Kölner Platz 1, 80804, Munich, Germany.

339) Institute of Clinical Chemistry and Pathobiochemistry, Klinikum rechts der Isar, Technical University of Munich, Munich, Germany.

340) Institute of Virology, School of Medicine, Technical University of Munich/Helmholtz Zentrum München, 81675 Munich, Germany; DZIF, partner sites Munich and Cologne/Bonn, Germany.

341) Institute of Medical Genetics and Applied Genomics, The University of Tübingen, Germany; NGS Competence Center Tübingen, The University of Tübingen, Germany.

342) Institute of Medical Genetics and Applied Genomics, University of Tübingen, Tübingen, Germany.

343) Institute of Medical Genetics and Applied Genomics, University of Tübingen, Calwerstrasse 7, 72076, Tübingen, Germany.

**-Quebec COVID-19 Biobank (BQC-19)**

J. Brent Richards^12,14,15^, Guillaume Butler-Laporte^12,13^

**-POLCOVID GEnomika**

Mirosław Kwasniewski^344^, Urszula Korotko^344^, Karolina Chwialkowska^344^, Magdalena Niemira^345^, Jerzy Jaroszewicz^346^, Barbara Sobala-Szczygiel^346^, Beata Puzanowska^347^, Anna Parfieniuk-Kowerda^348^, Diana Martonik^348^, Anna Moniuszko-Malinowska^349^, Sławomir Pancewicz^349^, Dorota Zarębska-Michaluk^350^, Krzysztof Simon^351^,Monika Pazgan-Simon^351^, Iwona Mozer-Lisewska^352^, Maciej Bura^352^, Agnieszka Adamek^352^, Krzysztof Tomasiewicz^353^ Małgorzata Pawłowska^354^, Anna Piekarska^355^, Aleksandra Berkan-Kawinska^355^, Andrzej Horban^356^, Justyna Kowalska^356^, Regina Podlasin^357^, Piotr Wasilewski^357^, Arsalin Azzadin^358^, Miroslaw Czuczwar^359^, Slawomir Czaban^360^, Paweł Olszewski^361^, Jacek Bogocz^361^, Magdalena Ochab^361^, Anna Kruk^361^, Sandra Uszok^361^, Agnieszka Bielska^361^, Anna Szałkowska^362^, Justyna Raczkowska^362^, Gabriela Sokołowska^362^ Joanna Chorostowska-Wynimko^363^, Aleksandra Jezela-Stanek^363^, Adriana Roży^363^, Urszula Lechowicz^363^, Urszula Polowianiuk^364^, Kamil Grubczak^365^, Aleksandra Starosz^365^, Andrzej Eljaszewicz^365^, Wiktoria Izdebska^365^, Adam Krętowski^367^, Robert Flisiak^368^, Marcin Moniuszko^369^

344) Centre for Bioinformatics and Data Analysis, Medical University of Bialystok, Bialystok, Poland.

345) Clinical Research Centre, Medical University of Bialystok, Bialystok, Poland.

346) Department of Infectious Diseases in Bytom, Medical University of Silesia, Silesia, Poland

347) Department of Infectious Diseases, Megrez Hospital in Tychy, Tychy, Poland

348) Department of Infectious Diseases and Hepatology, Medical University of Bialystok, Bialystok, Poland

349) Department of Infectious Diseases and Neuroinfection, Medical University of Bialystok, Biastolyk, Poland

350) Department of Infectious Diseases, Jan Kochanowski University, Kielce, Poland

351) Department of Infectious Diseases and Hepatology, Wroclaw Medical University, Wrocław, Poland

352) Department of Infectious Diseases and Hepatology, Poznan University of Medical Sciences, Poznan, Poland

353) Department of Infectious Diseases, Medical University of Lublin, Lublin, Poland

354) Department of Infectious Diseases and Hepatology, Nicolaus Copernicus University, Toruń, Poland

355) Department of Infectious Diseases and Hepatology, Medical University of Lodz, Lodz, Poland

356) Department of Adult Infectious Diseases, Warsaw Medical University, Warsaw, Poland

357) IV-th Department, Hospital for Infectious Diseases, Poland

358) District Hospital in Bielsk Podlaski, Poland

359) Department of Anaesthesiology and Intensive Therapy, Medical University of Lublin, Lublin, Poland

360) Department of Anaesthesiology and Intensive Therapy, Medical University of Bialystok, Bialystok, Poland

361) IMAGENE.ME SA, Bialystok, Poland

362) Clinical Research Centre, Medical University of Bialystok, Biastolyk, Poland

363) Department of Genetics and Clinical Immunology, National Institute of Tuberculosis and Lung Diseases in Warsaw, Warsaw, Poland

364) District Sanitary Inspectorate in Bialystok, Biastolyk, Poland

365) Department of Regenerative Medicine and Immune Regulation, Medical University of Bialystok, Biastolyk, Poland

366) Department of Allergology and Internal Medicine, Medical University of Bialystok, Bialystok, Poland

367) Clinical Research Centre, Medical University of Bialystok, Bialystok, Poland

368) Department of Infectious Diseases and Hepatology, Medical University of Bialystok, Bialystok, Poland

369) Department of Allergology and Internal Medicine; Department of Regenerative Medicine and Immune Regulation, Medical University of Bialystok, Bialystok, Poland

**- Saudi GENOME**

Malak Abedalthagafi^370^, Manal Alaamery^371,372,373^, Salam Massadeh^371,372,373^, Mohamed Fawzy^370^, Hadeel AlBardis^370^, Nora Aljawini^371,372^, Moneera Alsuwailm^371^, Faisal Almalki^371^, Serghei Mangul^374^, Junghyun Jung^374^

370) Genomics Research Department, Saudi Human Genome Project, King Fahad Medical City and King Abdulaziz City for Science and Technology, Riyadh, Saudi Arabia

371) Developmental Medicine Department, King Abdullah International Medical Research Center, King Saud Bin Abdulaziz University for Health Sciences, King Abdulaziz Medical City, Ministry of National Guard-Health Affairs

372) KACST-BWH Centre of Excellence for Biomedicine, Joint Centers of Excellence Program, King Abdulaziz City for Science and Technology (KACST), Riyadh, Saudi Arabia.

373) King Abdulaziz City for Science and Technology (KACST)-Saudi Human Genome Satellite Lab at Abdulaziz Medical City, Ministry of National Guard Health Affairs (MNGHA), Riyadh, Saudi Arabia.

374) Department of Clinical Pharmacy, School of Pharmacy, University of Southern California, Los Angeles, California, USA..

**- QATAR GENOME**

Hamdi Mbarek^375^, Chadi Saad^375^, Yaser Al-Sarraj^375^, Wadha Al-Muftah^375^, Radja Badji^375^, Asma Al Thani^375^, Said I. Ismail^375^

375) Qatar Genome Program, Qatar Foundation Research, Development and Innovation, Qatar Foundation, Doha, Qatar

