## Supplementary figures and images for "Common, low-frequency, rare, and ultra-rare coding variants contribute to COVID-19 severity"

### Supplementary Fig. 1

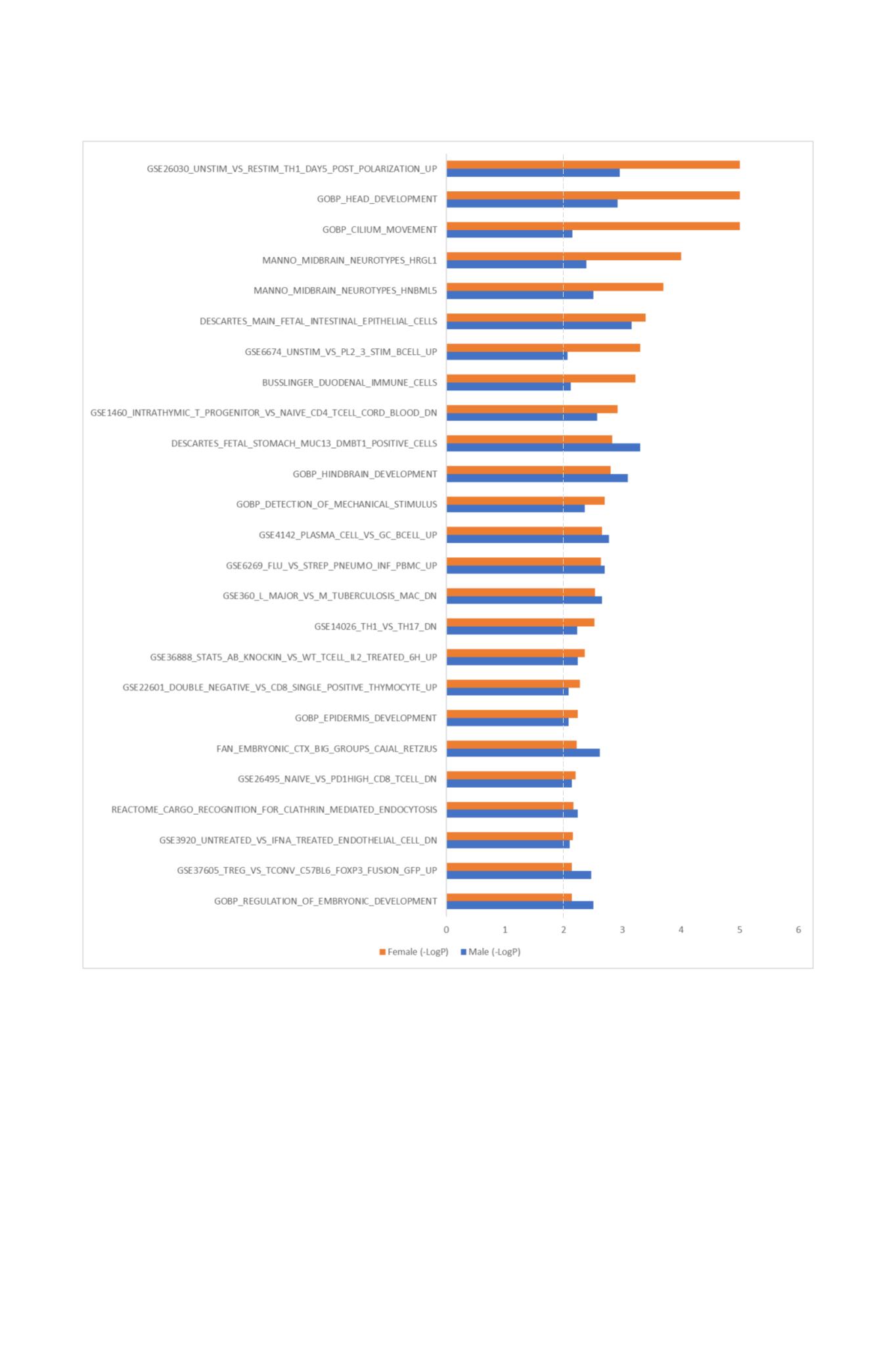

### Supplementary Fig. 2

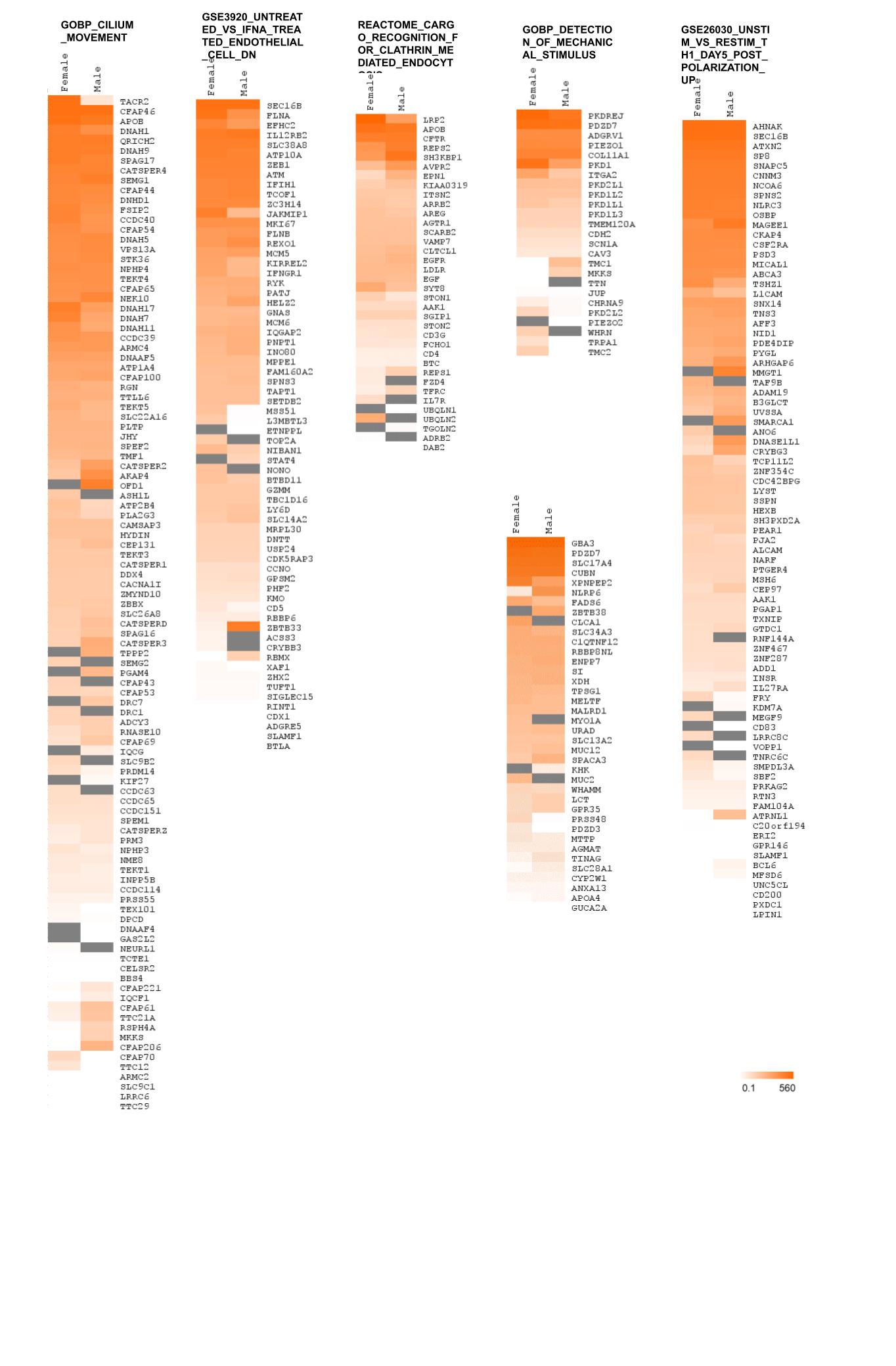

### Supplementary Fig. 3

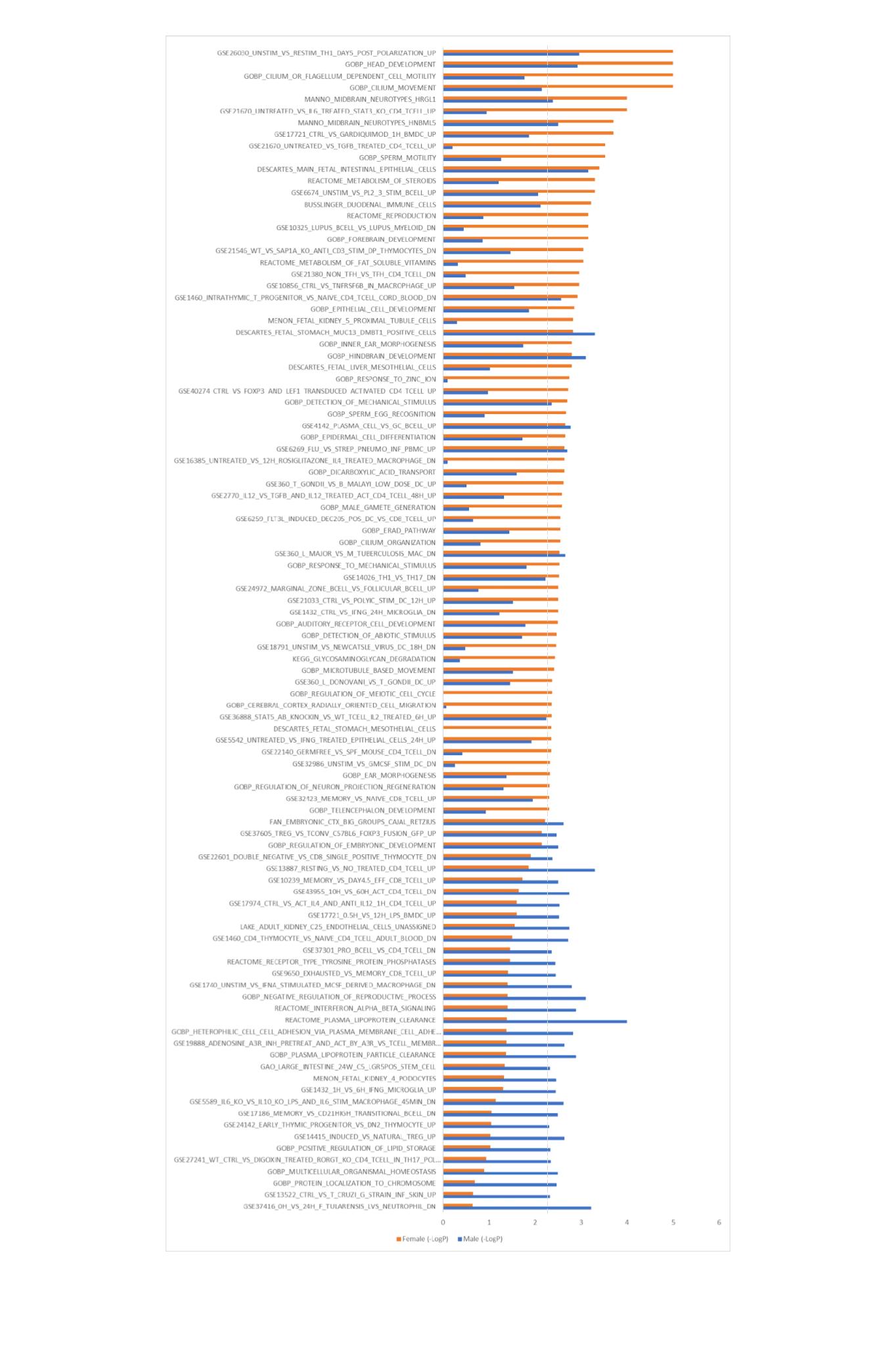

### Supplementary Fig. 4

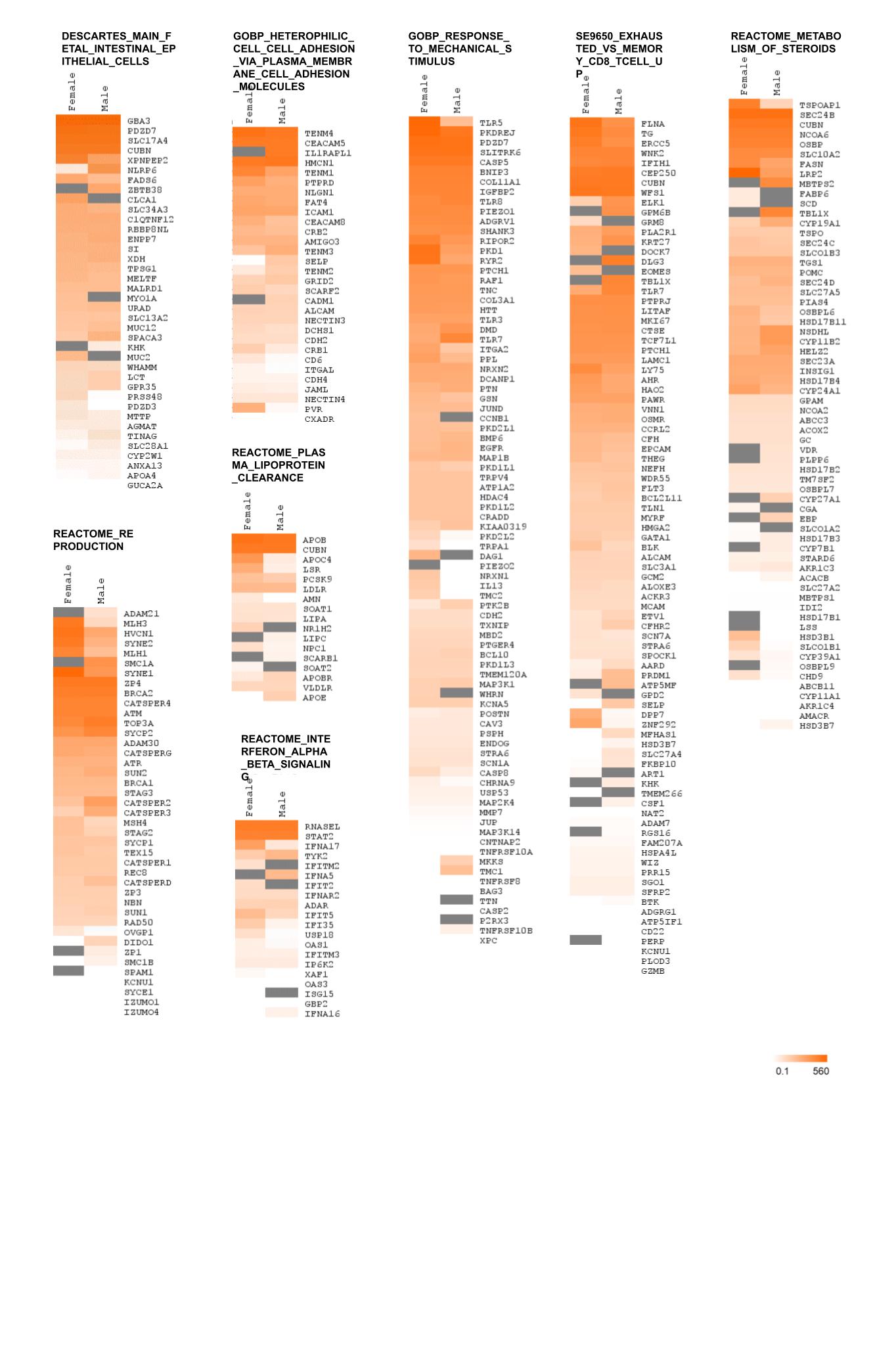
